# Left Atrial Remodeling, Hemodynamic Burden, and Time-Varying Risk of Newly Documented Atrial Fibrillation in Heart Failure With Preserved Ejection Fraction

**DOI:** 10.64898/2026.07.20.26358547

**Authors:** Lingyu Mi, Jeffrey Shi Kai Chan, Wing Tak Wong, Gary Tse, Fang Fang

## Abstract

**Background:** Left atrial volume index (LAVI) reflects chronic left atrial remodelling in heart failure with preserved ejection fraction (HFpEF), but its nonlinear, haemodynamic, and time-varying associations with newly documented atrial fibrillation (AF) remain uncertain.

**Methods:** We studied 764 patients with HFpEF without documented AF at baseline from a deidentified electronic health record registry in Hong Kong. Baseline LAVI was evaluated continuously, by tertiles, and using restricted cubic splines. Documented incident AF was defined as the first qualifying AF diagnosis or electrocardiographic record after the index echocardiogram. Cause-specific Cox and Fine–Gray models were used, with death before AF treated as a competing event. A haemodynamic overlap-adjusted model additionally included E/e′ ratio and pulmonary artery systolic pressure (PASP).

**Results:** During a median follow-up of 5.81 years, 360 patients developed documented incident AF and 272 died before AF was documented. In the primary clinical model, each 10-mL/m² increase in LAVI was associated with higher risk of documented incident AF in cause-specific Cox regression (HR, 1.08; 95% CI, 1.06–1.11) and Fine–Gray regression (sHR, 1.06; 95% CI, 1.04–1.09). After adjustment for E/e′ ratio and PASP, the continuous association was attenuated. However, the highest LAVI tertile remained associated with documented incident AF in both cause-specific Cox regression (HR, 1.78; 95% CI, 1.28–2.49) and Fine–Gray regression (sHR, 1.47; 95% CI, 1.05–2.04). Restricted cubic splines demonstrated a nonlinear association, with excess risk concentrated in the upper LAVI range. Period-specific analyses showed that the LAVI–AF association was strongest during the first year after echocardiography and attenuated thereafter.

**Conclusions:** In HFpEF patients without documented AF at baseline, marked left atrial enlargement identified a high-risk structural–haemodynamic phenotype for clinically documented incident AF. Routine echocardiographic measures may support risk-enriched rhythm surveillance, particularly early after echocardiographic assessment.

## Introduction

Heart failure with preserved ejection fraction (HFpEF) is a heterogeneous clinical syndrome^1^ in which atrial fibrillation (AF) is both common and clinically consequential^2,3^. Once AF is clinically documented, loss of coordinated atrial contraction, irregular ventricular activation, and rapid ventricular rates may further compromise ventricular filling and aggravate symptoms and haemodynamic congestion^4^. Conversely, elevated filling pressures, atrial stretch, and structural remodelling characteristic of HFpEF provide a substrate for electrical instability and AF susceptibility^1,2^. This bidirectional relationship is associated with greater symptom burden, more frequent heart failure hospitalisation, and adverse clinical outcomes^2,3^. Nevertheless, among patients with HFpEF without documented AF at baseline, the routinely measurable structural and haemodynamic features that identify those at greatest risk of subsequently documented AF remain incompletely defined^5^.

The left atrium reflects the cumulative effects of chronic pressure and volume loading in HFpEF^6,7^. Left atrial volume index (LAVI) is a routinely available measure of left atrial remodelling and integrates the long-term consequences of elevated left ventricular filling pressure, atrial stretch, and structural adaptation^6^. However, left atrial enlargement does not occur in isolation. It is closely related to contemporaneous haemodynamic abnormalities, including an elevated ratio of early transmitral flow velocity to early diastolic mitral annular velocity (E/e′) and increased pulmonary artery systolic pressure (PASP), which reflect left ventricular filling pressure and downstream pulmonary circulatory load, respectively^6–8^. Accordingly, LAVI may capture both an accumulated atrial substrate and the haemodynamic burden underlying its development^8,9^. Because LAVI, E/e′, and PASP are routinely reported during clinical echocardiography, their combined interpretation may offer a practical means of characterising AF susceptibility in HFpEF without requiring advanced imaging techniques^7,10^.

Prior studies have established left atrial enlargement as an important marker of AF risk^10,11^, and haemodynamic abnormalities have also been linked to AF susceptibility^8,10,12^. However, several clinically relevant uncertainties remain in HFpEF. First, it is unclear whether the association between LAVI and subsequently documented AF is linear across the full range of left atrial size or instead concentrated among patients with more advanced left atrial enlargement^10,12^. Second, the extent to which LAVI provides risk information that overlaps with contemporaneous filling-pressure and pulmonary haemodynamic burden remains uncertain^8,10^. Third, prior risk-prediction analyses have generally evaluated baseline echocardiographic measures as fixed predictors of subsequent AF, although a single baseline examination may be more informative for near-term AF documentation than for events recorded many years later^10,12^. Finally, because death is frequent in older patients with HFpEF and may preclude the observation of clinically documented incident AF, competing risk needs to be considered when estimating both cumulative incidence and associations^13^.

Clarifying these issues has practical relevance because routine echocardiography may help identify patients with HFpEF who could benefit from more targeted rhythm surveillance^5,10^. We therefore investigated the association between baseline LAVI and clinically documented incident AF in a longitudinal cohort of patients with HFpEF without documented AF at baseline. To place left atrial remodelling within a broader physiological context, we additionally examined routinely available echocardiographic measures of filling pressure and pulmonary haemodynamic load, including E/e′ ratio and PASP^6^. We hypothesised that marked left atrial enlargement would identify a high-risk structural–haemodynamic phenotype for clinically documented incident AF, and that this excess risk would be most pronounced during the early years after baseline echocardiography.

## Methods

### Study design and population

This retrospective cohort study used a real-world registry of patients with heart failure with preserved ejection fraction (HFpEF) derived from deidentified electronic health records in Hong Kong. Clinical information and longitudinal outcomes were obtained through the Clinical Data Analysis and Reporting System (CDARS), an integrated electronic health record system that captures inpatient and outpatient diagnoses, investigations, treatment records, and clinical outcomes across public healthcare facilities in Hong Kong^14,15^.

This study used an anonymised dataset that is publicly documented in a dissertation accessible via ProQuest (Tse G. Atrial cardiomyopathy and heart failure, 2023; https://www.proquest.com/docview/3252768154). The submission to ProQuest was managed and approved by the Graduate School as part of the graduation requirements. The study originally received approval from the Joint Chinese University of Hong Kong–New Territories East Cluster Clinical Research Ethics Committee (approval number 2019.422) as part of the doctoral studies of G.T.. The requirement for individual informed consent was waived by the Committee owing to the retrospective nature of the study. The study was conducted in accordance with the Declaration of Helsinki and reported in accordance with the Strengthening the Reporting of Observational Studies in Epidemiology guidelines.

Adults aged 18 years or older with a physician-documented diagnosis of HFpEF between 1 January 2010 and 31 December 2016 were eligible. HFpEF was defined as a left ventricular ejection fraction (LVEF) of at least 50% on echocardiography together with objective clinical or echocardiographic evidence consistent with contemporary diagnostic recommendations^16,17^. Patients with identifiable hypertrophic, restrictive, or infiltrative cardiomyopathy and those with identifiable severe valvular heart disease were excluded when these conditions were ascertainable from clinical records and/or diagnostic coding.

The index date was defined as the date of the baseline echocardiographic examination. When more than one eligible examination was available, the first qualifying echocardiogram was selected. Patients with documented pre-existing atrial fibrillation (AF) on or before the index date were excluded.

### Clinical and echocardiographic assessment

Baseline demographic characteristics, heart rate, comorbidities, and medication use were obtained from structured electronic health records. Prespecified clinical variables included age, sex, diabetes mellitus, renal disease, hypertension, chronic obstructive pulmonary disease, and ischaemic heart disease. Baseline beta-blocker and diuretic use were additionally considered in a treatment-adjusted sensitivity analysis. Covariates were selected a priori according to their clinical relevance to HFpEF, left atrial remodelling, and AF susceptibility, rather than on the basis of univariable statistical significance.

Comprehensive transthoracic echocardiography was performed by experienced echocardiographers using standard imaging views, and measurements were obtained and reported in accordance with contemporary echocardiographic recommendations. Echocardiographic variables were extracted from structured clinical echocardiography reports generated at the index examination. No de novo core-laboratory remeasurement was performed for the present study.

The extracted variables included LVEF, left ventricular mass index, relative wall thickness, left ventricular end-diastolic diameter, left atrial diameter, the left atrial-to-aortic root ratio, left atrial volume index (LAVI), the ratio of early transmitral flow velocity to early diastolic mitral annular velocity (E/e′), and pulmonary artery systolic pressure (PASP). Left ventricular mass and left atrial volume were indexed to body surface area according to guideline-based reporting conventions. The reported E/e′ ratio was used as an estimate of left ventricular filling pressure, whereas PASP was considered a measure of downstream pulmonary haemodynamic load.

The primary exposure was baseline LAVI. LAVI was evaluated as a continuous variable per 10-mL/m^2^ increase, according to tertiles, and as a nonlinear continuous exposure. Tertile cut points were derived from the observed pre-imputation distribution and fixed before model fitting: T1, ≤42.27 mL/m^2^; T2, >42.27 to ≤61.69 mL/m^2^; and T3, >61.69 mL/m^2^. The same cut points were applied throughout the primary and sensitivity analyses.

To place left atrial remodelling within a broader physiological context, LAVI, E/e′ ratio, and PASP were additionally evaluated as routinely available echocardiographic phenotypes representing left atrial structure, left ventricular filling pressure, and pulmonary haemodynamic load, respectively. Each phenotype was standardised using its standard deviation in the observed pre-imputation distribution and assessed in a separate multivariable model; no formal between-marker comparison was performed.

### Ascertainment of atrial fibrillation and follow-up

Pre-existing AF was defined as any documented diagnosis of AF occurring on or before the index echocardiography date. All available medical history before the index date was searched using inpatient and outpatient records in CDARS, based on prespecified diagnostic codes and available electrocardiographic records. Because systematic rhythm monitoring was not performed, baseline AF status was defined according to documented diagnoses and available electrocardiographic records in routine clinical care.

Incident AF was defined as the first qualifying AF diagnosis or electrocardiographic record after the index date. The earliest qualifying inpatient, outpatient, or available electrocardiographic record was used as the event date. AF ascertainment was primarily based on routinely recorded diagnostic information in CDARS and was supplemented by available electrocardiographic records. Accordingly, incident AF in this study represents clinically documented AF newly recorded after the index echocardiogram, rather than systematically detected AF onset.

Patients were followed from the index date until documented incident AF, death, the last available clinical record, or the administrative end of follow-up on 31 December 2019, whichever occurred first. Death before documented incident AF was treated as a competing event. Patients who remained alive and free from documented AF at their last available record or at the administrative end of follow-up were censored.

Follow-up time was calculated from the index date to documented incident AF, competing death, or censoring. The incidence rate of documented incident AF was calculated by dividing the number of incident events by the accumulated person-time at risk and was expressed per 1,000 person-years with a 95% confidence interval.

### Statistical analysis

Continuous variables are presented as mean with standard deviation or median with interquartile range, as appropriate, and categorical variables as number and percentage. Baseline characteristics according to LAVI tertiles were described among patients with observed pre-imputation LAVI measurements. Approximately normally distributed continuous variables were compared using one-way analysis of variance, non-normally distributed continuous variables using the Kruskal–Wallis test, and categorical variables using the Pearson χ^2^ test or Fisher’s exact test, as appropriate. These comparisons were descriptive, and no adjustment for multiple testing was applied.

Cumulative incidence functions were used to estimate the probability of documented incident AF while accounting for death before AF as a competing event. Differences across observed LAVI tertiles were assessed using Gray’s test, and cumulative incidence estimates with 95% confidence intervals were calculated at prespecified time points^18^.

Missing baseline covariates were handled using multiple imputation by chained equations, with 30 imputed datasets generated^19^. The imputation model included the variables used in the primary and sensitivity analyses, competing-risk outcome status, and follow-up time. Derived LAVI variables were reconstructed within each completed dataset using the prespecified definitions. Statistical models were fitted separately in each imputed dataset, and estimates and covariance matrices were combined using Rubin’s rules^19^.

The association between LAVI and documented incident AF was assessed using complementary cause-specific Cox proportional hazards and Fine–Gray subdistribution hazards regression[26858290]. In the cause-specific Cox models, death before AF was treated as a censoring event, whereas in the Fine–Gray models it was retained as a competing event. Results are reported as hazard ratios (HRs) or subdistribution hazard ratios (sHRs) with 95% confidence intervals.

Four progressively adjusted models were prespecified. Model 1 included age and sex, and Model 2 additionally included heart rate. Model 3, designated as the primary clinical model, further adjusted for LVEF, left ventricular mass index, diabetes mellitus, renal disease, hypertension, chronic obstructive pulmonary disease, and ischaemic heart disease. Model 4 was defined as the haemodynamic overlap-adjusted model and additionally included E/e′ ratio and PASP. This model was used to evaluate whether the association between LAVI and documented incident AF shared risk information with contemporaneous filling-pressure and pulmonary haemodynamic burden. It was not intended to establish formal mediation, causal pathways, or etiologic independence.

For tertile analyses, T1 served as the reference category. Overall P values were obtained from joint 2-degree-of-freedom Wald tests of the T2 and T3 coefficients, and P values for trend were obtained by modelling tertile rank as an ordinal variable.

Restricted cubic spline models were used to examine potential nonlinearity in the association between LAVI and documented incident AF^20^. The proportional hazards assumption was assessed using scaled Schoenfeld residuals^21^. To characterise potential variation in the magnitude of association over follow-up, period-specific cause-specific Cox and Fine–Gray models were fitted for 0–1 year, greater than 1–3 years, and greater than 3 years^22^.

Sensitivity analyses included complete-case analyses; landmark analyses at 30 days, 90 days, and 1 year; analyses using left atrial diameter and the left atrial-to-aortic root ratio as alternative measures of left atrial size; analyses addressing extreme LAVI values; and a treatment-adjusted model additionally including baseline beta-blocker and diuretic use. Detailed specifications for the imputation, nonlinear, time-varying, and sensitivity analyses are provided in the **Supplementary Methods**.

All tests were 2-sided, and P<0.05 was considered statistically significant. Analyses were performed using R software (version 4.6.0).

## Results

### Study population and clinical outcomes

The source cohort comprised 792 patients with heart failure with preserved ejection fraction (HFpEF) who underwent baseline echocardiography. After exclusion of 28 patients with documented atrial fibrillation (AF) on or before the index examination, 764 patients without documented AF at baseline were included in the present analysis (**Figure 1**). The median follow-up was 5.81 years (IQR, 2.28–9.34). During 4108.1 person-years of follow-up, clinically documented incident AF was recorded in 360 patients; 272 patients died before AF was documented, and 132 patients remained event-free or were censored. The incidence rate of documented incident AF was 87.6 per 1,000 person-years (95% CI, 78.8–97.2).

**Figure 1.**
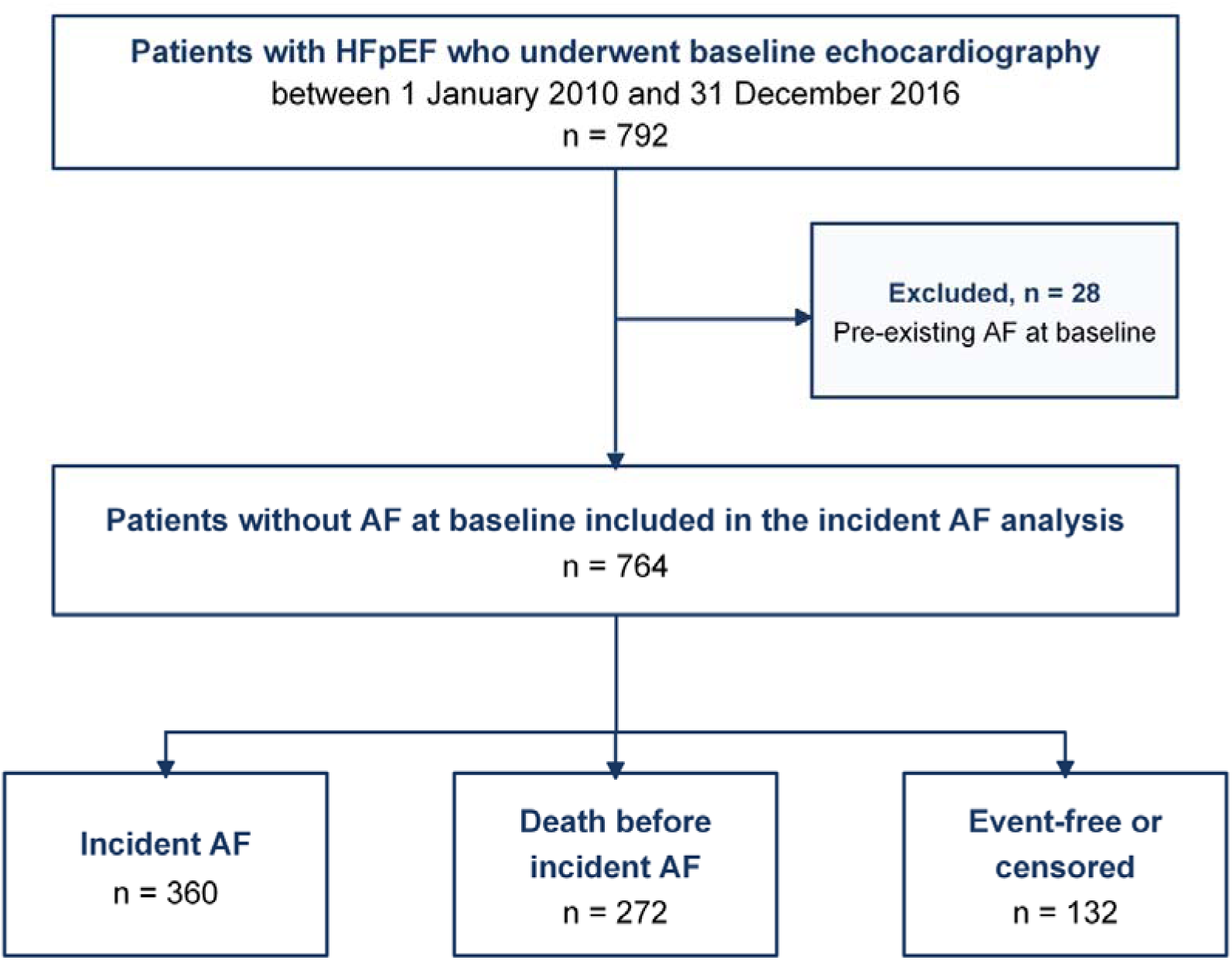
Study Population Selection and Competing-Risk Outcome Classification. Flow diagram of patient selection from the source HFpEF cohort to the final analytic cohort. After exclusion of patients with documented AF on or before the index echocardiogram, 764 patients without documented AF at baseline were included in the present analysis. During follow-up, 360 patients had documented incident AF, 272 died before AF was documented, and 132 remained event-free or were censored. Baseline LAVI was available in 715 patients and missing in 49 patients; primary multivariable analyses used the full cohort after multiple imputation. Documented incident AF was defined as the first qualifying AF diagnosis or electrocardiographic record after the index echocardiogram in routine care. AF, atrial fibrillation; HFpEF, heart failure with preserved ejection fraction; LAVI, left atrial volume index.

Baseline left atrial volume index (LAVI) was available in 715 patients and missing in 49 patients. Demographic characteristics, heart rate, comorbidities, and baseline medication variables were complete, whereas missingness among echocardiographic variables ranged from 0.4% for left ventricular ejection fraction (LVEF) to 8.0% for the left atrial-to-aortic root ratio (**Supplementary Table 2**).

### Baseline characteristics according to LAVI

Among patients with observed baseline LAVI, the mean age was 75.95±11.36 years, 322 patients (45.0%) were male, and the median LAVI was 43.00 mL/m^2^ (IQR, 40.99–67.81 mL/m^2^). Median LAVI was 38.95 mL/m^2^ in T1, 43.01 mL/m^2^ in T2, and 86.91 mL/m^2^ in T3 (**Table 1**).

**Table 1.** Baseline Characteristics According to Tertiles of Left Atrial Volume Index.

| Characteristic | Overall<br>N = 715 | T1 ($\leq 42.27$ mL/m<sup>2</sup>)<br>N = 239 | T2 ($>42.27$ to $\leq 61.69$ mL/m<sup>2</sup>)<br>N = 238 | T3 ($>61.69$ mL/m<sup>2</sup>)<br>N = 238 | P value |
| --- | --- | --- | --- | --- | --- |
| **Demographic and clinical characteristics** |  |  |  |  |  |
| Age, years | 75.95 ± 11.36 | 77.12 ± 8.25 | 74.95 ± 11.42 | 75.77 ± 13.69 | 0.107 |
| Male sex | 322 (45.0%) | 148 (61.9%) | 97 (40.8%) | 77 (32.4%) | <0.001 |
| Heart rate, beats/min | 79.06 ± 20.33 | 73.76 ± 16.39 | 77.55 ± 21.15 | 85.89 ± 21.23 | <0.001 |
| **Comorbidities** |  |  |  |  |  |
| Diabetes mellitus | 228 (31.9%) | 75 (31.4%) | 78 (32.8%) | 75 (31.5%) | 0.948 |
| Renal disease | 94 (13.1%) | 40 (16.7%) | 29 (12.2%) | 25 (10.5%) | 0.116 |
| Hypertension | 388 (54.3%) | 132 (55.2%) | 131 (55.0%) | 125 (52.5%) | 0.808 |
| Chronic obstructive pulmonary disease | 115 (16.1%) | 44 (18.4%) | 37 (15.5%) | 34 (14.3%) | 0.456 |
| Ischaemic heart disease | 323 (45.2%) | 110 (46.0%) | 109 (45.8%) | 104 (43.7%) | 0.862 |
| **Medications** |  |  |  |  |  |
| Beta-blocker use | 367 (51.3%) | 120 (50.2%) | 119 (50.0%) | 128 (53.8%) | 0.654 |
| Diuretic use | 364 (50.9%) | 109 (45.6%) | 127 (53.4%) | 128 (53.8%) | 0.134 |
| **Echocardiographic characteristics** |  |  |  |  |  |
| Left ventricular ejection fraction, % | 60.97 ± 7.95 | 62.91 ± 8.35 | 60.36 ± 7.56 | 59.60 ± 7.56 | <0.001 |
| Left ventricular mass index, g/m<sup>2</sup> | 102.60 (85.04–143.91) | 87.43 (76.84–115.04) | 92.18 (83.34–145.70) | 122.78 (103.01–156.32) | <0.001 |
| Relative wall thickness | 0.51 (0.43–0.56) | 0.53 (0.44–0.60) | 0.51 (0.42–0.54) | 0.50 (0.44–0.54) | <0.001 |
| Left ventricular end-diastolic diameter, cm | 3.96 (3.63–4.99) | 3.80 (3.61–4.64) | 3.80 (3.63–5.11) | 4.40 (3.68–5.00) | <0.001 |
| Left atrial diameter, cm | 3.49 (3.33–3.79) | 3.33 (3.19–3.44) | 3.51 (3.36–3.73) | 3.80 (3.52–4.15) | <0.001 |
| Left atrial-to-aortic root ratio | 1.42 (1.28–1.60) | 1.26 (1.21–1.33) | 1.46 (1.39–1.57) | 1.61 (1.48–1.89) | <0.001 |
| Left atrial volume index, mL/m<sup>2</sup> | 43.00 (40.99–67.81) | 38.95 (36.87–41.01) | 43.01 (42.64–53.27) | 86.91 (67.81–123.45) | — |

| Characteristic | Overall<br>N = 715 | T1 ($\leq 42.27$ mL/m<sup>2</sup>)<br>N = 239 | T2 ($> 42.27$ to $\leq 61.69$ mL/m<sup>2</sup>)<br>N = 238 | T3 ($> 61.69$ mL/m<sup>2</sup>)<br>N = 238 | P value |
| --- | --- | --- | --- | --- | --- |
| E/e' ratio | 18.92 (16.32–21.66) | 16.54 (13.98–18.44) | 19.03 (17.54–20.85) | 21.34 (19.02–23.94) | <0.001 |
| Pulmonary artery<br>systolic pressure, mm<br>Hg | 39.77 (34.64–49.17) | 35.95 (33.92–39.81) | 38.89 (34.05–47.00) | 49.50 (40.00–56.30) | <0.001 |

Values are presented as mean ± standard deviation, median (interquartile range), or n (%), as appropriate. P values were calculated using one-way analysis of variance for approximately normally distributed continuous variables, the Kruskal–Wallis test for non-normally distributed continuous variables, and the Pearson $\chi^2$ test or Fisher's exact test for categorical variables, as appropriate. The analysis included patients with observed pre-imputation left atrial volume index measurements; missing values were not imputed for descriptive presentation. No P value was calculated for left atrial volume index because it defined the grouping variable. COPD, chronic obstructive pulmonary disease; IHD, ischaemic heart disease; LAVI, left atrial volume index; LVEF, left ventricular ejection fraction; PASP, pulmonary artery systolic pressure.

Age and the prevalences of diabetes mellitus, renal disease, hypertension, chronic obstructive pulmonary disease, and ischaemic heart disease were similar across LAVI tertiles. Baseline beta-blocker and diuretic use also did not differ significantly. In contrast, male sex was less frequent across increasing LAVI tertiles, decreasing from 61.9% in T1 to 32.4% in T3, whereas mean heart rate increased from 73.76 to 85.89 beats/min.

Increasing LAVI was accompanied by a broader echocardiographic phenotype of left atrial enlargement and greater haemodynamic burden. Median left atrial diameter increased from 3.33 cm in T1 to 3.80 cm in T3, and the median left atrial-to-aortic root ratio increased from 1.26 to 1.61. Median E/e′ ratio increased from 16.54 to 21.34, and median pulmonary artery systolic pressure increased from 35.95 to 49.50 mm Hg. In parallel, left ventricular mass index was higher in T3 than in T1, and left ventricular end-diastolic diameter was also greater in T3, whereas relative wall thickness was modestly lower across increasing LAVI tertiles. Mean LVEF was modestly lower in T3 than in T1.

When patients were classified according to competing-risk outcome status, patients with subsequently documented incident AF had higher baseline LAVI, left atrial diameter, left atrial-to-aortic root ratio, E/e′ ratio, and PASP than those who remained event-free and those who died before AF was documented (**Supplementary Table 1**). The death-before-AF group was older, whereas the documented incident AF group showed the most prominent left atrial and haemodynamic abnormalities.

### Cumulative incidence of documented incident AF

The cumulative incidence of documented incident AF differed significantly across LAVI tertiles, with early separation of the T3 curve from the lower tertiles (Gray’s test P<0.001; Figure 2). In the overall cohort, the cumulative incidence of documented incident AF was 7.3% (95% CI, 5.5–9.2) at 1 year, 17.8% (95% CI, 15.1–20.6) at 3 years, and 26.7% (95% CI, 23.6–29.9) at 5 years. At 5 years, the cumulative incidence increased from 15.6% (95% CI, 11.0–20.3) in T1 and 19.9% (95% CI, 14.8–25.0) in T2 to 43.4% (95% CI, 37.1–49.8) in T3; corresponding event counts and competing death counts are shown in **Supplementary Table 3**.

**Figure 2.**
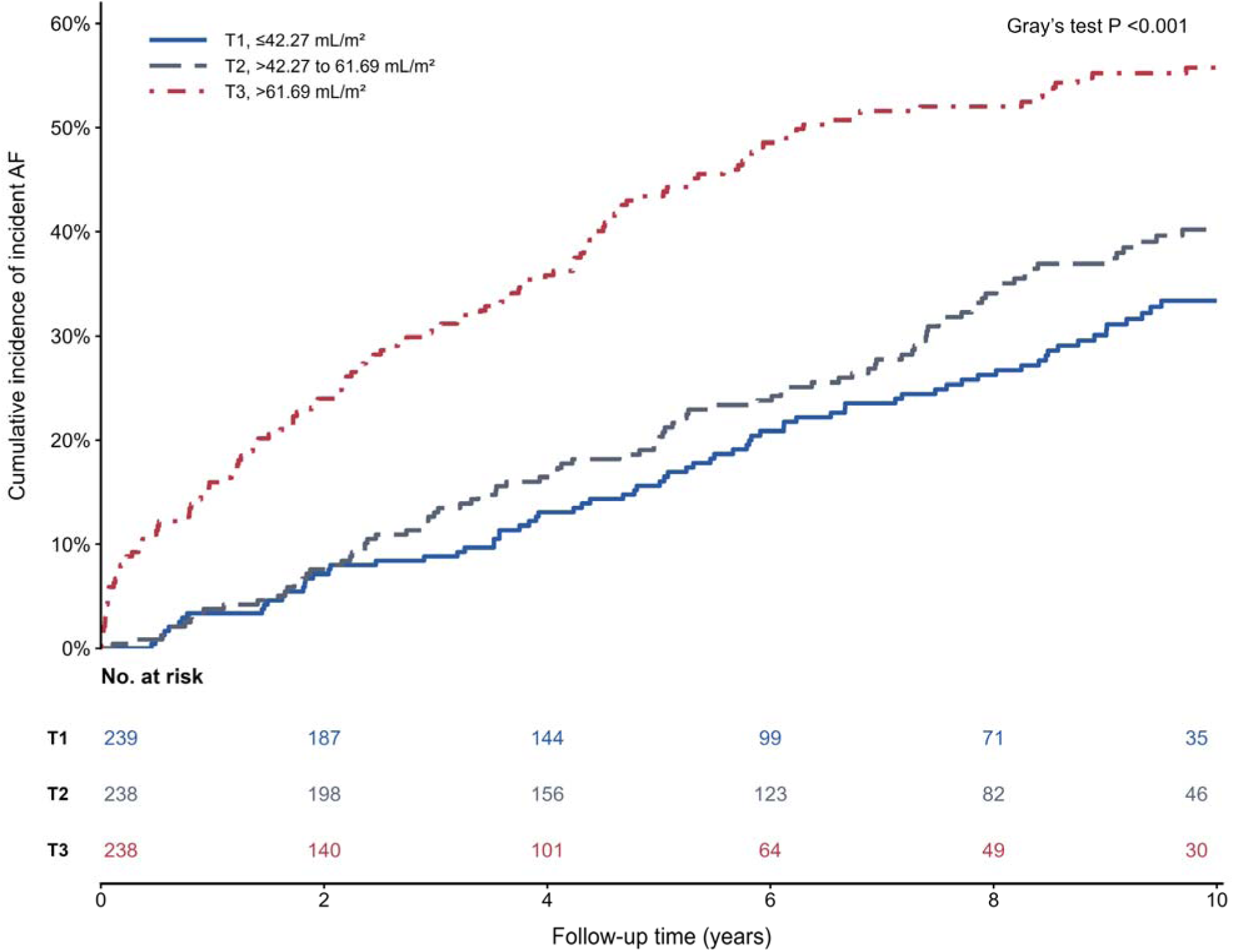
Cumulative Incidence of Documented Incident Atrial Fibrillation According to Tertiles of Left Atrial Volume Index. Cumulative incidence functions for documented incident AF are shown according to tertiles of left atrial volume index: T1, ≤42.27 mL/m²; T2, >42.27 to ≤61.69 mL/m²; and T3, >61.69 mL/m². The analysis included 715 patients with available pre-imputation LAVI measurements. Documented incident AF was defined as the first qualifying AF diagnosis or electrocardiographic record after the index echocardiogram among patients without documented AF at baseline. Death before documented incident AF was treated as a competing event, and differences among groups were assessed using Gray’s test over all available follow-up. Curves are displayed through 10 years for graphical clarity; all available follow-up was retained in the regression analyses. Numbers at risk are shown below the plot. AF, atrial fibrillation; LAVI, left atrial volume index.

### Overall association of LAVI with documented incident AF

In progressively adjusted models, higher LAVI was associated with greater risk of documented incident AF before inclusion of contemporaneous haemodynamic measures (**Table 2**). In the primary clinical model, each 10-mL/m^2^ increase in LAVI was associated with a higher cause-specific hazard of documented incident AF (HR, 1.08; 95% CI, 1.06–1.11; P<0.001) and a higher subdistribution hazard after accounting for competing death (sHR, 1.06; 95% CI, 1.04–1.09; P<0.001). In the haemodynamic overlap-adjusted Model 4, which additionally included E/e′ ratio and PASP, the continuous association was attenuated in both the cause-specific Cox model (HR, 1.03; 95% CI, 0.99–1.06; P=0.106) and the Fine–Gray model (sHR, 1.02; 95% CI, 0.98–1.05; P=0.377).

**Table 2.**
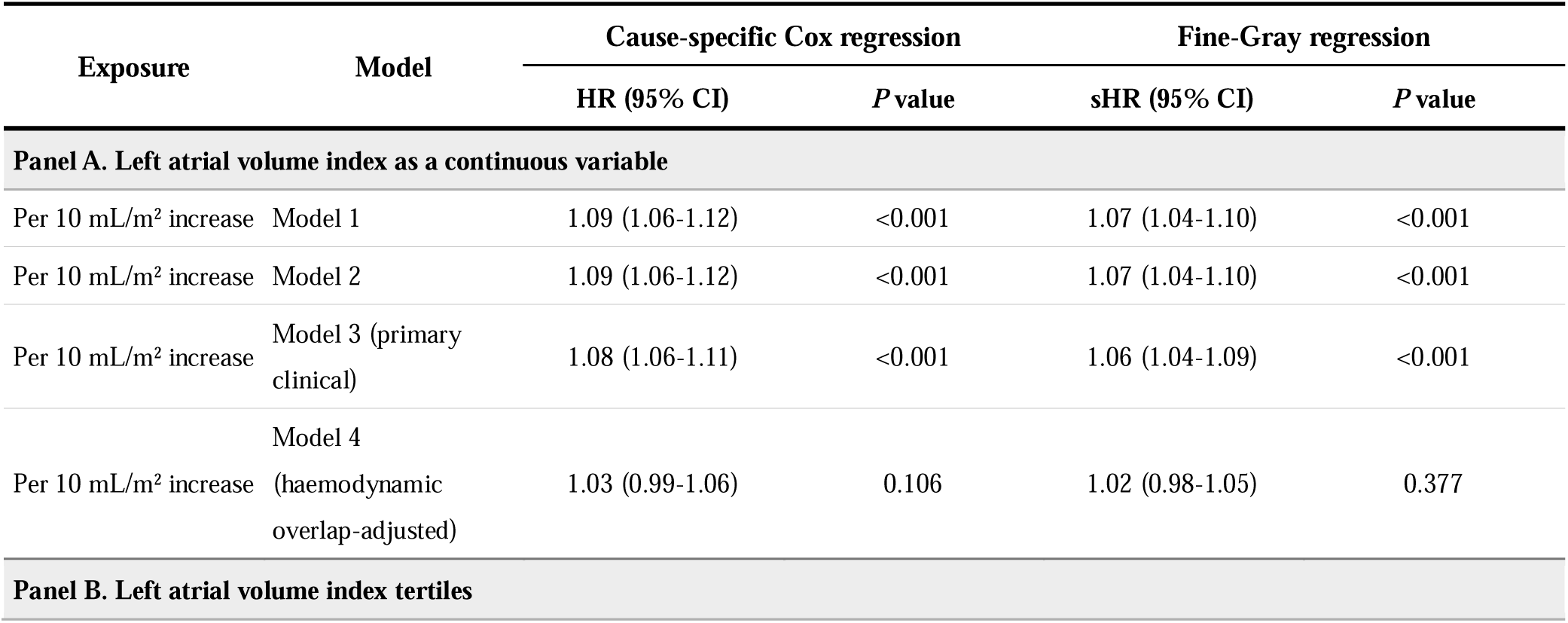
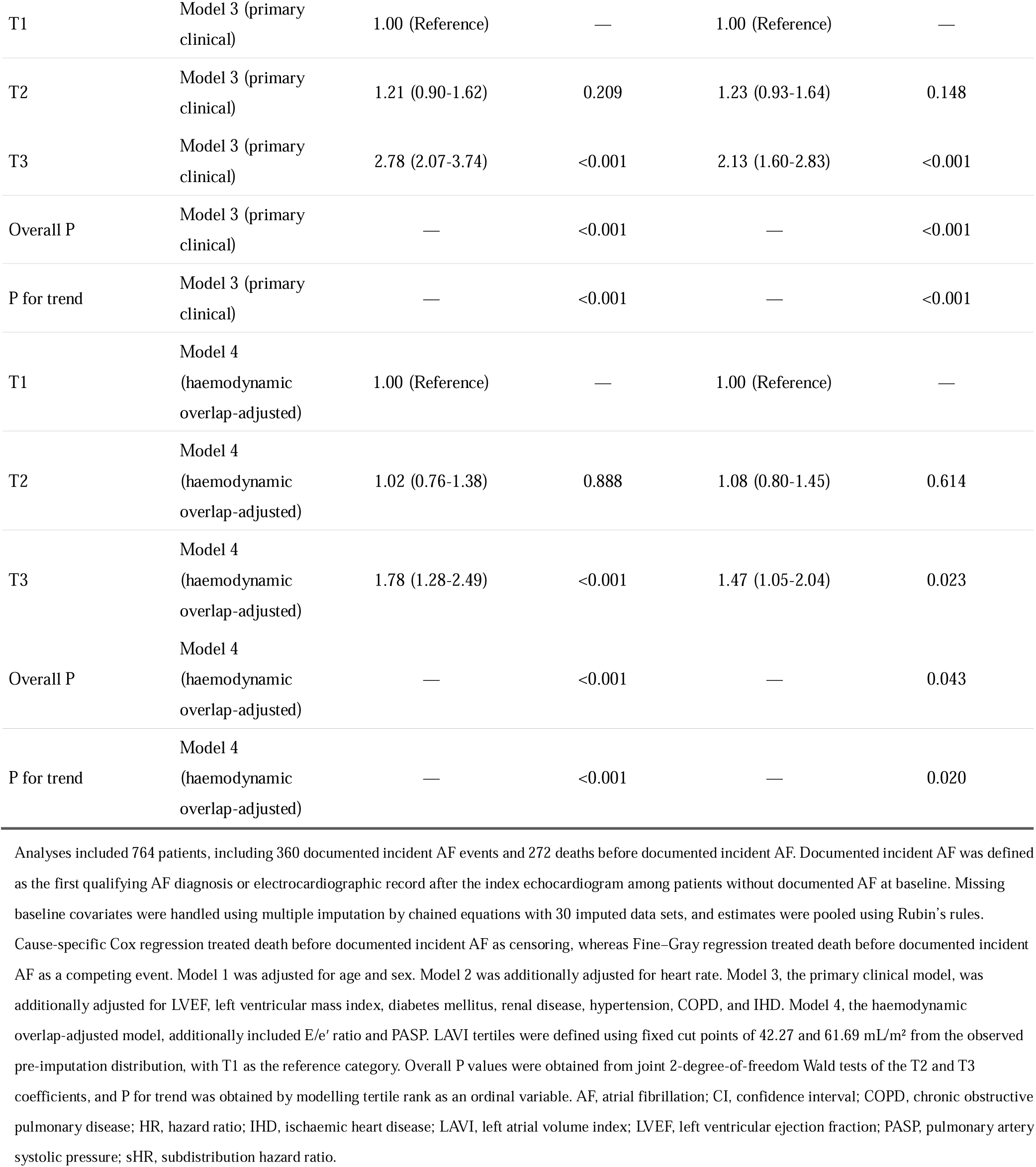
Associations of Left Atrial Volume Index With Documented Incident Atrial Fibrillation After Multiple Imputation.

The tertile analyses showed a threshold-like pattern. Compared with T1, T2 was not significantly associated with documented incident AF in either the primary clinical model or the haemodynamic overlap-adjusted Model 4. By contrast, T3 was associated with substantially higher risk in the primary clinical model, with an HR of 2.78 (95% CI, 2.07–3.74; P<0.001) and an sHR of 2.13 (95% CI, 1.60–2.83; P<0.001). In the haemodynamic overlap-adjusted Model 4, the association for T3 remained significant in both the cause-specific Cox model (HR, 1.78; 95% CI, 1.28–2.49; P<0.001) and the Fine–Gray model (sHR, 1.47; 95% CI, 1.05–2.04; P=0.023). Overall tertile associations remained significant in both models.

### Nonlinear association and echocardiographic phenotypes

Restricted cubic spline analyses in the primary clinical model demonstrated significant overall associations between LAVI and documented incident AF in both cause-specific Cox and Fine–Gray regression (both overall P<0.001; **Figure 3**). The association was nonlinear in the cause-specific analysis (P for nonlinearity <0.001) and also showed evidence of nonlinearity in the Fine–Gray analysis (P for nonlinearity=0.014). The estimated risk rose more prominently across the upper range of LAVI and flattened at the most extreme values, where the confidence intervals widened.

**Figure 3.**
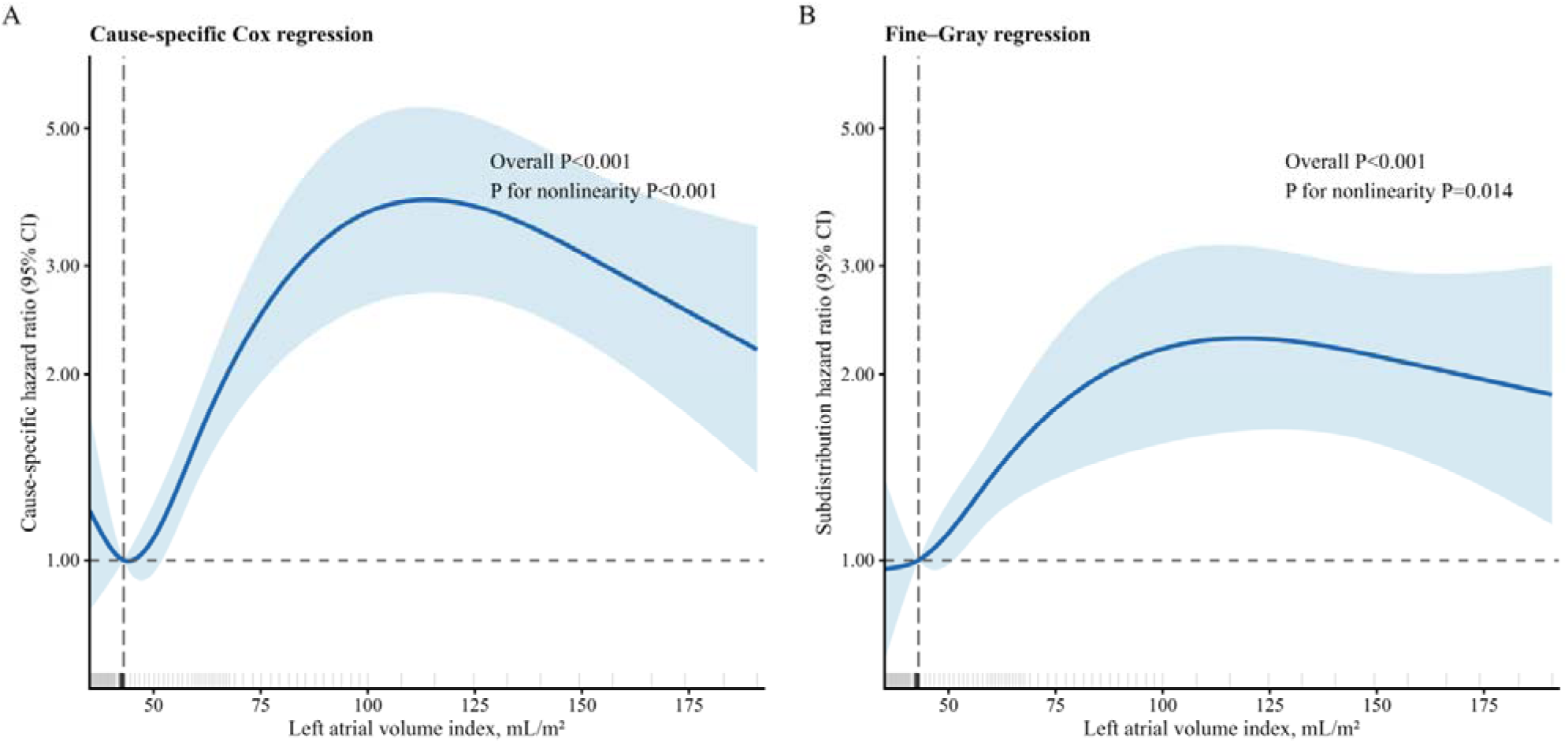
Restricted Cubic Spline Associations of Left Atrial Volume Index With Documented Incident Atrial Fibrillation in the Primary Clinical Model. Restricted cubic spline analyses illustrating the association between left atrial volume index and documented incident AF using cause-specific Cox regression (A) and Fine–Gray regression (B). Both analyses were based on Model 3, the primary clinical adjustment model, which included age, sex, heart rate, left ventricular ejection fraction, left ventricular mass index, diabetes mellitus, renal disease, hypertension, chronic obstructive pulmonary disease, and ischaemic heart disease. Solid lines represent the pooled effect estimates, and shaded areas indicate the corresponding 95% confidence intervals. The horizontal dashed line denotes an effect estimate of 1.00, and the vertical dashed line indicates the reference LAVI value of 43.0 mL/m². Rug marks along the x-axis show the distribution of observed LAVI values. Death before documented incident AF was treated as a censoring event in the cause-specific Cox model and as a competing event in the Fine–Gray model. Overall and nonlinear association P values are displayed in each panel. Missing covariate data were handled using multiple imputation with 30 imputed data sets, and estimates were pooled according to Rubin’s rules. AF, atrial fibrillation; CI, confidence interval; HR, hazard ratio; LAVI, left atrial volume index; sHR, subdistribution hazard ratio.

In the haemodynamic overlap-adjusted Model 4, the spline associations were attenuated, particularly in the competing-risk analysis, but the graphical pattern remained broadly consistent with a higher-risk upper range of LAVI (**Supplementary Figure 1**). Similar nonlinear patterns were observed after winsorisation of extreme LAVI values and after exclusion of observed values above the 99th percentile, supporting that the overall spline pattern was not driven solely by extreme observations (**Supplementary Figure 2**).

When routinely available echocardiographic phenotypes were evaluated on a standardised scale, LAVI, E/e′ ratio, and PASP were each associated with documented incident AF in separate primary clinical models (**Figure 4** and **Supplementary Table 4**). Per 1-SD increase, LAVI was associated with an HR of 1.33 (95% CI, 1.22–1.46) and an sHR of 1.25 (95% CI, 1.13–1.37). E/e′ ratio was associated with an HR of 1.26 (95% CI, 1.17–1.35) and an sHR of 1.20 (95% CI, 1.13–1.28). PASP was associated with an HR of 1.51 (95% CI, 1.39–1.64) and an sHR of 1.42 (95% CI, 1.26–1.60). All associations had P<0.001.

**Figure 4.**
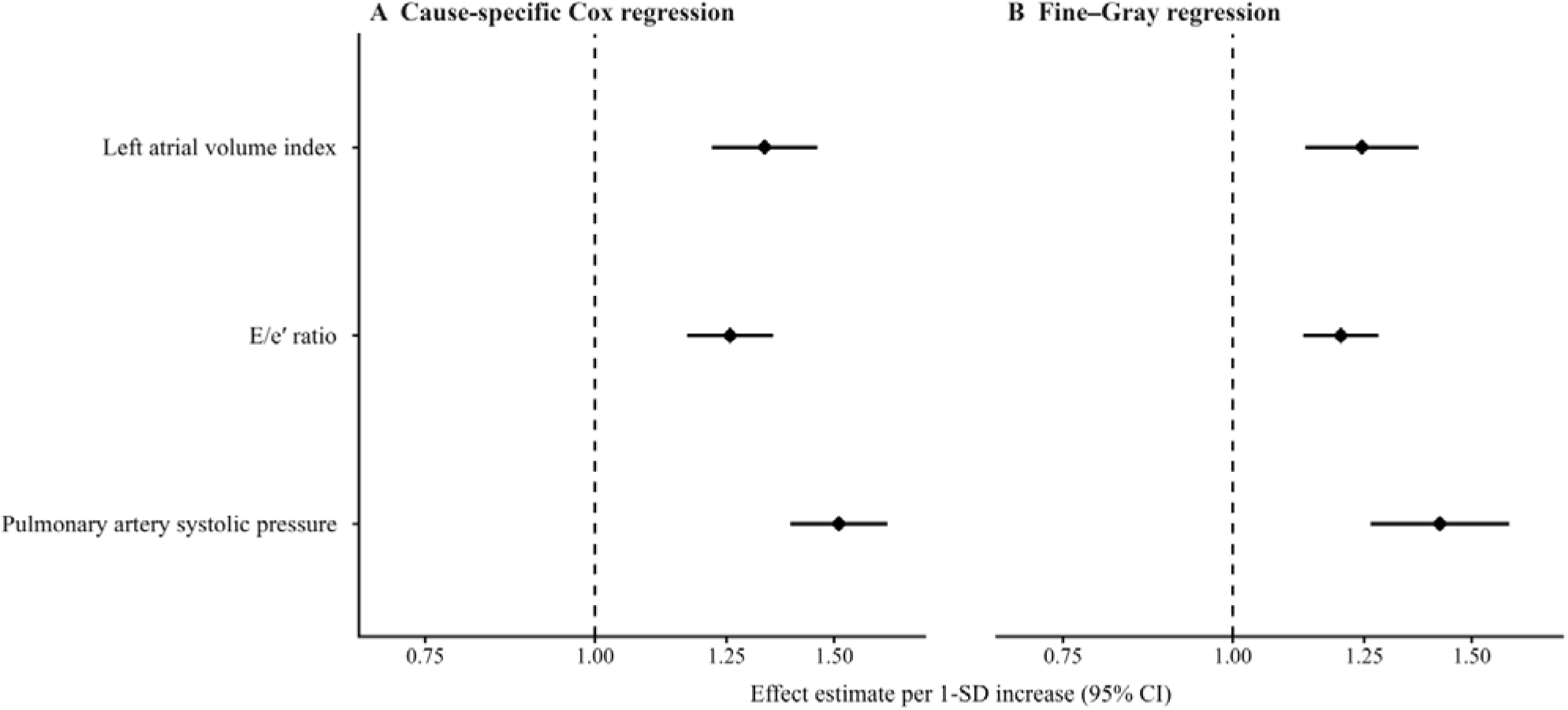
Adjusted Associations of Selected Echocardiographic Phenotypes With Documented Incident Atrial Fibrillation. Forest plots showing the associations of left atrial volume index, E/e′ ratio, and pulmonary artery systolic pressure with documented incident AF using cause-specific Cox regression (A) and Fine–Gray regression (B). Effect estimates are presented per 1-standard-deviation increase in each echocardiographic phenotype. Diamonds represent the pooled hazard ratios or subdistribution hazard ratios, and horizontal lines indicate the corresponding 95% confidence intervals. The vertical dashed line denotes an effect estimate of 1.00. Each phenotype was evaluated in a separate Model 3, the primary clinical adjustment model, which included age, sex, heart rate, left ventricular ejection fraction, left ventricular mass index, diabetes mellitus, renal disease, hypertension, chronic obstructive pulmonary disease, and ischaemic heart disease. Death before documented incident AF was treated as a censoring event in the cause-specific Cox model and as a competing event in the Fine–Gray model. Standard deviations were calculated from the observed pre-imputation distributions. Missing covariate data were handled using multiple imputation with 30 imputed data sets, and estimates were pooled according to Rubin’s rules. AF, atrial fibrillation; CI, confidence interval; E/e′, ratio of early transmitral flow velocity to early diastolic mitral annular velocity; HR, hazard ratio; LAVI, left atrial volume index; LV, left ventricular; PASP, pulmonary artery systolic pressure; sHR, subdistribution hazard ratio.

Correlation and variance inflation factor analyses showed correlations among left atrial structural and haemodynamic measures, but did not indicate substantial multicollinearity in the haemodynamic overlap-adjusted Model 4 (**Supplementary Figure 3**).

### Time-varying association of LAVI with documented incident AF

Scaled Schoenfeld residual testing showed evidence of nonproportionality for LAVI terms in the cause-specific Cox models, including continuous LAVI and the LAVI tertile contrasts (**Supplementary Table 10** and **Supplementary Figure 4**). Period-specific analyses showed that the magnitude of association with documented incident AF varied across follow-up intervals.

In the primary clinical model, the association between continuous LAVI and documented incident AF was strongest during the first year and decreased over time (**Table 3**). Per 10-mL/m^2^ increase, the cause-specific HR was 1.15 (95% CI, 1.10–1.20; P<0.001) during 0–1 year, 1.08 (95% CI, 1.03–1.13; P=0.002) during >1–3 years, and 1.06 (95% CI, 1.02 – 1.10; P=0.002) beyond 3 years (P for time interaction=0.004). Corresponding Fine–Gray estimates were 1.14 (95% CI, 1.09–1.19; P<0.001), 1.05 (95% CI, 1.00–1.11; P=0.043), and 1.04 (95% CI, 1.00–1.07; P=0.049), respectively (P for time interaction=0.002).

**Table 3.**
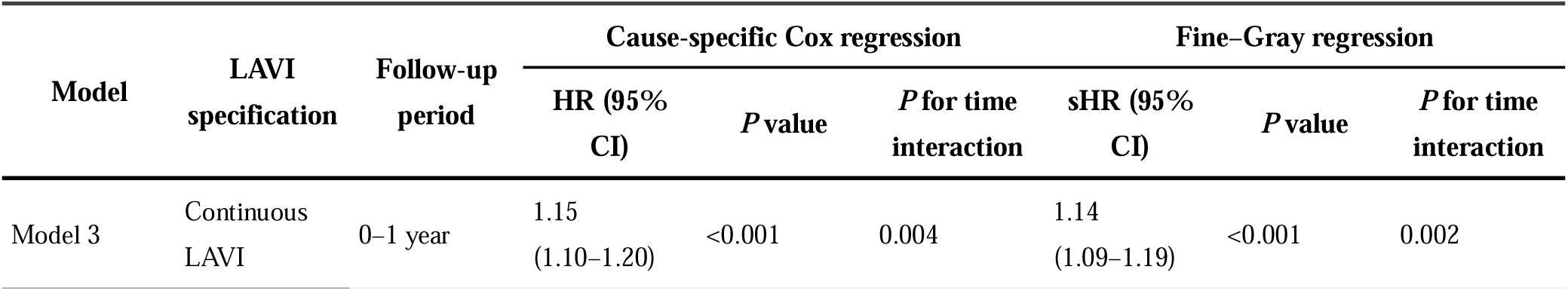
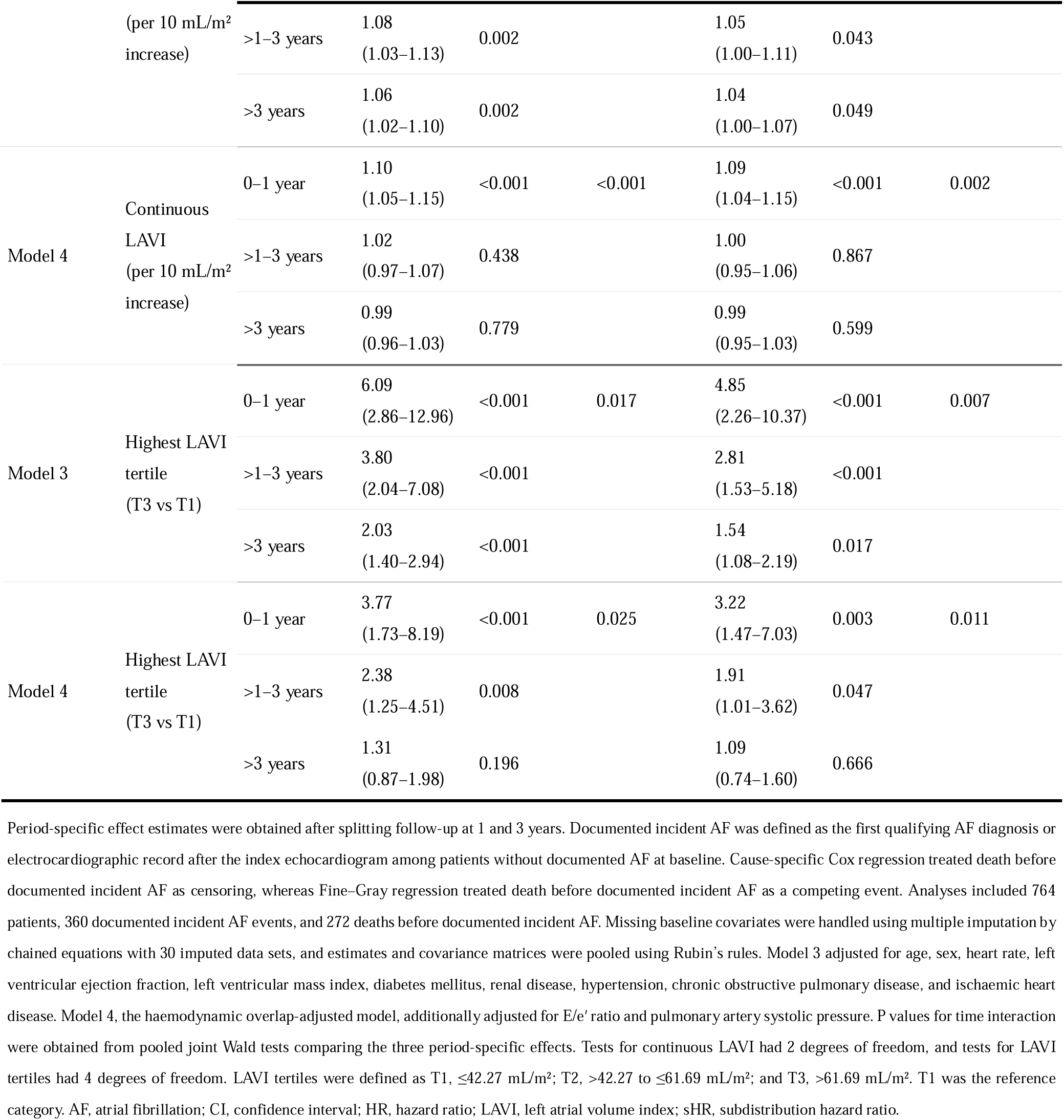
Period-Specific Associations of Left Atrial Volume Index With Documented Incident Atrial Fibrillation.

In the haemodynamic overlap-adjusted Model 4, the continuous LAVI association remained significant during the first year in both the cause-specific Cox model (HR, 1.10; 95% CI, 1.05–1.15; P<0.001) and the Fine–Gray model (sHR, 1.09; 95% CI, 1.04–1.15; P<0.001), but was no longer significant during later intervals. Time interactions remained significant in both models.

A similar time-varying pattern was observed for the highest LAVI tertile. In the primary clinical model, the cause-specific HRs for T3 versus T1 were 6.09 (95% CI, 2.86–12.96; P<0.001) during 0–1 year, 3.80 (95% CI, 2.04–7.08; P<0.001) during >1–3 years, and 2.03 (95% CI, 1.40–2.94; P<0.001) beyond 3 years (P for time interaction=0.017). The corresponding Fine–Gray estimates were 4.85 (95% CI, 2.26–10.37; P<0.001), 2.81 (95% CI, 1.53–5.18; P<0.001), and 1.54 (95% CI, 1.08–2.19; P=0.017), respectively (P for time interaction=0.007).

In the haemodynamic overlap-adjusted Model 4, T3 remained associated with documented incident AF during 0–1 year (HR, 3.77; 95% CI, 1.73–8.19; P<0.001; sHR, 3.22; 95% CI, 1.47–7.03; P=0.003) and >1–3 years (HR, 2.38; 95% CI, 1.25–4.51; P=0.008; sHR, 1.91; 95% CI, 1.01–3.62; P=0.047). Beyond 3 years, the estimates were attenuated and no longer statistically significant in either model. Detailed period-specific analyses, including T2 estimates and the number of patients entering each follow-up interval, are provided in **Supplementary Table 12**.

### Sensitivity analyses

The main findings were consistent in the complete-case analyses (**Supplementary Table 5**). In the haemodynamic overlap-adjusted Model 4, T3 remained associated with documented incident AF compared with T1 in both the cause-specific Cox analysis (HR, 1.79; 95% CI, 1.25–2.56; P=0.001) and the Fine–Gray analysis (sHR, 1.59; 95% CI, 1.12–2.27; P=0.010). In contrast, the continuous LAVI association was attenuated in the haemodynamic overlap-adjusted Model 4. Complete-case period-specific Cox analyses also showed a similar declining pattern over follow-up, with the strongest associations observed during the first year (**Supplementary Table 11**).

Landmark analyses at 30 days, 90 days, and 1 year produced a similar overall pattern (**Supplementary Table 6**). In the 30-day landmark cohort, T3 remained associated with subsequently documented AF in the haemodynamic overlap-adjusted Model 4 in both the cause-specific Cox model (HR, 1.62; 95% CI, 1.15– 2.28; P=0.006) and the Fine–Gray model (sHR, 1.56; 95% CI, 1.11–2.20; P=0.011). Corresponding estimates were also significant in the 90-day landmark analysis (HR, 1.57; 95% CI, 1.11–2.22; P=0.010; sHR, 1.64; 95% CI, 1.16–2.31; P=0.005) and in the 1-year landmark analysis (HR, 1.50; 95% CI, 1.03–2.17; P=0.034; sHR, 1.92; 95% CI, 1.32–2.78; P<0.001). The continuous LAVI association in Model 4 remained nonsignificant across landmark analyses.

Alternative measures of left atrial size were also associated with documented incident AF (**Supplementary Table 7**). Per 1-SD increase, left atrial diameter was associated with a higher cause-specific hazard (HR, 1.40; 95% CI, 1.28–1.53; P<0.001) and subdistribution hazard (sHR, 1.46; 95% CI, 1.29–1.64; P<0.001). The left atrial-to-aortic root ratio showed similar associations in the cause-specific Cox model (HR, 1.40; 95% CI, 1.27–1.54; P<0.001) and the Fine–Gray model (sHR, 1.53; 95% CI, 1.38–1.69; P<0.001).

Analyses addressing extreme LAVI values did not materially alter the overall interpretation (**Supplementary Table 8**). After winsorisation at the 1st and 99th percentiles, continuous LAVI remained associated with documented incident AF in the primary clinical model, whereas the haemodynamic overlap-adjusted Model 4 showed attenuation. After excluding observed LAVI values above the 99th percentile, the Model 3 association remained significant in both regression frameworks, and the Model 4 cause-specific association was also statistically significant, although the Fine–Gray association remained attenuated. The corresponding spline sensitivity analyses showed broadly similar nonlinear patterns **(Supplementary Figure 2).**

Additional adjustment for baseline beta-blocker and diuretic use yielded results comparable with the haemodynamic overlap-adjusted Model 4 (**Supplementary Table 9**). In this treatment-adjusted model, continuous LAVI was not significantly associated with documented incident AF, whereas T3 remained associated with higher risk in both the cause-specific Cox model (HR, 1.79; 95% CI, 1.28–2.49; P<0.001) and the Fine–Gray model (sHR, 1.46; 95% CI, 1.05–2.03; P=0.025).

Across sensitivity analyses, the association between marked left atrial enlargement and documented incident AF remained consistent, whereas intermediate LAVI elevation and continuous LAVI in the haemodynamic overlap-adjusted models showed weaker or nonsignificant associations. The time-varying analyses further indicated that the excess risk associated with high LAVI was most prominent during the early years after baseline echocardiography.

## Discussion

In this retrospective cohort study of patients with HFpEF without documented AF at baseline, newly documented AF and death before AF were both frequent, supporting the use of a competing-risk framework. LAVI was associated with subsequent AF in a nonlinear and time-varying manner, with excess risk concentrated among patients with marked LA enlargement and greatest during the early years after baseline echocardiography. The attenuation of the continuous LAVI association after inclusion of E/e′ ratio and PASP suggests that LA remodelling and haemodynamic burden represent overlapping dimensions of the HFpEF substrate, whereas the persistent risk associated with the highest LAVI tertile indicates that advanced LA enlargement retains clinically relevant information beyond contemporaneous haemodynamic measures.

Prior community-based studies established LA volume as a predictor of incident AF in individuals without prevalent AF and showed that LA volume provides risk information beyond conventional clinical factors and linear LA dimension^11^. More recent population-based and HFpEF studies have further linked LA structure and function, including LA strain, with subsequent AF risk^10,23^. However, those cohorts were not uniformly enriched for HFpEF and have not routinely examined whether the association was nonlinear, time-varying, or influenced by the high competing mortality burden characteristic of older HFpEF populations^10,11,23^. Our study extends this evidence by demonstrating that, in a real-world HFpEF cohort, the risk associated with LAVI was not uniformly distributed across the full range of LA size but was concentrated in patients with marked LA enlargement. The highest LAVI tertile had more than twice the cause-specific hazard of newly documented AF in the primary clinical model, and this association persisted after adjustment for E/e′ ratio and PASP. These findings suggest that advanced LA enlargement in HFpEF reflects more than chamber dilatation alone; it identifies a clinically relevant atrial substrate embedded within a broader disease phenotype^9,10^.

Our findings also complement prior HFpEF studies emphasizing the importance of LA structure and function^10,24,25^. TOPCAT-based analyses and other HFpEF cohorts have shown that LA enlargement, LA dysfunction, and impaired LA strain are common and prognostically relevant in HFpEF^10,24,25^, and studies using more intensive rhythm surveillance or advanced atrial imaging have linked LA remodelling and dysfunction with AF development^5,10^. The present study adds to this literature by focusing on routinely reported echocardiographic measures, evaluating LAVI together with E/e′ ratio and PASP, accounting for death before AF as a competing event, and demonstrating that the LAVI–AF association was strongest early after echocardiography. Thus, our findings do not challenge the incremental value of LA strain or advanced atrial imaging; rather, they suggest that conventional echocardiographic markers, when interpreted together, can identify a high-risk phenotype associated with newly documented AF in HFpEF.

The attenuation of the continuous LAVI association in the haemodynamic overlap-adjusted Model 4 should be interpreted in this context. LAVI reflects the cumulative structural consequences of chronic atrial pressure and volume loading, whereas E/e′ ratio and PASP represent contemporaneous filling-pressure and downstream pulmonary vascular load^6,8^. These measures are biologically interrelated in HFpEF; therefore, adding E/e′ ratio and PASP may account for shared risk information rather than negate the clinical relevance of LAVI^6,8,9^. The persistence of the highest LAVI tertile after haemodynamic overlap adjustment suggests that advanced LA enlargement retains risk information beyond these contemporaneous measures.

This interpretation also explains why the tertile and spline analyses remained clinically informative even when the continuous association was attenuated. A purely linear interpretation would imply a constant risk increase across the LAVI distribution, whereas our findings suggest that excess risk was concentrated in the upper range. The middle tertile was not significantly associated with AF compared with the lowest tertile, while the highest tertile showed consistently elevated risk. In HFpEF, marked LA enlargement may therefore represent an advanced atrial-myopathy-like stage that integrates chronic loading, interstitial remodelling, and increasing electrical vulnerability^9,10,26^. The consistency of this pattern across complete-case, landmark, treatment-adjusted, and extreme-value analyses supports its robustness.

The phenotype analyses further showed that LAVI, E/e′ ratio, and PASP each carried AF-related risk information when evaluated on a standardised scale. Numerically, PASP showed the largest association, followed by LAVI and E/e′ ratio, although these markers were assessed in separate models and were not formally compared. PASP may capture a more advanced haemodynamic state, integrating elevated left-sided filling pressure, pulmonary vascular load, right-sided consequences, recurrent congestion, and greater clinical contact during follow-up^8,27^. Thus, AF susceptibility in HFpEF appears to reflect the interaction between atrial remodelling, elevated LV filling pressure, and pulmonary vascular or right-sided haemodynamic burden^6,8,9,27^. The variance inflation factor analysis did not suggest substantial multicollinearity in the haemodynamic overlap-adjusted Model 4, supporting the interpretation that attenuation of the LAVI association reflects overlapping pathophysiological information rather than a purely statistical artefact.

A particularly novel aspect of this study is the time-varying association between LAVI and newly documented AF. The association was strongest during the first year after baseline echocardiography and attenuated thereafter, particularly after accounting for E/e′ ratio and PASP. These findings suggest that baseline LAVI is most informative for near- to intermediate-term AF documentation rather than serving as a fixed long-term risk marker. This pattern is clinically plausible because a single echocardiographic examination may best reflect the current atrial substrate and haemodynamic state^10^, whereas AF documentation over longer follow-up is increasingly influenced by intervening clinical events, treatment changes, healthcare contact, and competing death^5,13,28^.

Several mechanisms may explain this temporal pattern. Marked LA enlargement may identify patients with an advanced arrhythmogenic substrate or previously unrecognised paroxysmal AF, leading to earlier clinical documentation^9^. This is particularly relevant in an EHR-based cohort in which the low prevalence of documented baseline AF should not be interpreted as a low true AF burden in HFpEF^5,28^. The persistence of associations in 30-day, 90-day, and 1-year landmark analyses argues against the findings being explained solely by very early AF detection, but high LAVI may still identify patients closer to the threshold for clinical recognition. Over longer follow-up, incident AF documentation may be increasingly shaped by intervening clinical events, comorbidity progression, treatment changes, repeated decompensation, healthcare contact, and competing death^5,13,28^. Therefore, the declining association should be interpreted as attenuation of the information carried by a single baseline echocardiogram in a dynamic clinical syndrome, rather than disappearance of the biological relevance of LA enlargement.

Differences between our findings and studies reporting more persistent or linearly increasing risks may partly reflect differences in cohort structure, follow-up duration, and AF ascertainment^5,10,12,28^. Some prior studies included younger or more selected patients, incorporated more intensive rhythm surveillance, or focused on advanced imaging such as LA strain^5,10,12^. In contrast, our cohort was older and based on real-world electronic health records, which primarily captured clinically recognised AF events over routine follow-up. These differences are not simply limitations; they emphasise that AF risk markers may perform differently depending on whether the endpoint is systematically detected AF, newly documented AF in routine care, or longer-term AF burden^5,28^.

The clinical implications are practical but should remain within appropriate boundaries. LAVI, E/e′ ratio, and PASP are widely available during routine echocardiography and do not require advanced imaging software^6^. In patients with HFpEF without documented baseline AF, marked LA enlargement, particularly when accompanied by elevated E/e′ ratio or PASP, may identify a group in whom a single baseline ECG is insufficient to exclude clinically relevant AF susceptibility^5,10^. These patients may be considered for repeated 12-lead ECG assessment, ambulatory rhythm monitoring, review of rhythm data from implanted devices when available, or closer rhythm-focused follow-up, especially during the first year after echocardiography^5,29^. Importantly, these findings support risk-enriched rhythm surveillance rather than treatment allocation. LAVI should therefore be viewed as a marker to enrich rhythm-surveillance strategies, not as a stand-alone basis for anticoagulation or rhythm-control decisions^29^.

Several strengths should be noted. The study included a relatively large real-world HFpEF cohort with longitudinal follow-up and a large number of documented incident AF events. The analyses accounted for death before AF as a competing event, which is essential in an older HFpEF population with substantial mortality. LAVI was evaluated as a continuous variable, according to tertiles, and using restricted cubic splines, allowing us to identify a threshold-like and nonlinear risk pattern. The inclusion of E/e′ ratio and PASP helped place LA remodelling within a broader haemodynamic framework, while time-varying analyses addressed the important assumption that baseline echocardiographic risk markers have constant associations with documented AF over follow-up. The consistency of the findings across complete-case, landmark, alternative LA-measure, extreme-value, and treatment-adjusted analyses further supports the robustness of the main conclusions.

This study has several limitations. First, the retrospective design precludes causal inference. The associations among LAVI, haemodynamic burden, and newly documented AF should therefore be interpreted as risk-marker relationships rather than evidence that LA enlargement directly causes AF. The haemodynamic overlap-adjusted Model 4 was intended to assess shared risk information among interrelated echocardiographic measures, not to provide a formal mediation analysis. Second, AF ascertainment relied on routinely recorded diagnoses supplemented by available electrocardiographic records, without systematic prospective rhythm monitoring or central ECG adjudication. Asymptomatic or short paroxysmal AF episodes may therefore have been missed before and after the index echocardiogram^5,28^. Accordingly, the proportion of patients with documented AF at baseline should not be assumed to represent the true AF burden in HFpEF, and the present findings should be interpreted as identifying risk markers for AF newly documented during routine care rather than the true biological onset of AF^5,28^. Third, echocardiographic variables were extracted from structured clinical reports rather than core-laboratory remeasurement, which may have introduced measurement variability; however, this approach also reflects how these parameters are used in real-world practice. Fourth, LA strain, LA function, natriuretic peptides, obesity, sleep-disordered breathing, and longitudinal changes in echocardiographic parameters were not consistently available, limiting assessment of whether functional atrial measures would improve risk characterisation beyond LAVI and haemodynamic variables^10,30^. Fifth, although patients with identifiable severe valvular heart disease were excluded during source cohort construction, detailed information on valvular lesion type and severity was not consistently available in the analytic dataset; residual confounding by less severe or incompletely documented valvular disease cannot be excluded. Finally, this was a single-region cohort from Hong Kong, and external validation in other populations and healthcare systems is needed to determine the generalisability of these findings and their potential role in guiding AF surveillance strategies in HFpEF.

## Data Availability

This study used a dataset hosted on ProQuest Dissertations & Theses Global (ProQuest document view: 3252768154). The dataset originated from the doctoral thesis work of Dr Gary Tse at the Graduate School, The Chinese University of Hong Kong, and submission to ProQuest was managed and approved by the Graduate School as part of the graduation requirements. Access to the dataset is subject to the availability and terms of the third-party host platform.

## Conclusions

Among patients with HFpEF without documented AF at baseline, marked left atrial enlargement identified a structural–haemodynamic phenotype associated with higher risk of clinically documented incident AF, particularly during the early years after baseline echocardiography. Attenuation of the continuous LAVI association after adjustment for E/e′ ratio and PASP suggests substantial overlap between atrial remodelling and contemporaneous haemodynamic burden, whereas the persistent risk associated with the highest LAVI tertile supports the clinical relevance of advanced LA enlargement. These findings support the use of routinely available echocardiographic measures for risk-enriched rhythm surveillance in HFpEF, while highlighting the need to consider competing death and time-varying risk when interpreting long-term AF risk.

## Declarations

### Ethics approval and consent to participate

Ethics approval covering the source dataset and related secondary analyses was obtained from the Chinese University of Hong Kong–New Territories East Cluster Clinical Research Ethics Committee (approval number: 2019.422). The study was conducted in accordance with the Declaration of Helsinki. The requirement for informed consent was waived by the Ethics Committee owing to the retrospective use of the data and the absence of patient contact. Ethics approvals are publicly accessible via the CUHK–NTEC CREC approved-study database (search term: “tse gary”).

## Consent for publication

All authors consent to the publication of this manuscript.

## Funding

This research was supported by the Clinical and Translational Medicine Research Projects of CAMS (2023-I2M-C&T-B-057); National High Level Hospital Clinical Research Funding (2023-GSP-GG-34); and the Key Technology Research and Device Development Project for Innovative Diagnosis and Treatment of Structural Heart Disease in the Southwest Plateau Region (202302AA310045).

## Competing interests

None declared

## Acknowledgements

Not applicable.

## Author contributions

Lingyu Mi contributed to study conception and design, statistical analysis, data interpretation, figure and table preparation, and drafting of the manuscript. Jeffrey Shi Kai Chan contributed to methodology, statistical analysis, data validation, and critical revision of the manuscript. Wing Tak Wong contributed to data curation, study resources, methodological support, and critical revision of the manuscript. Gary Tse contributed to study conception, data resources, methodology, supervision, and critical revision of the manuscript. Fang Fang contributed to study conception and design, supervision, project administration, funding acquisition, and critical revision of the manuscript. All authors contributed to interpretation of the findings, reviewed and approved the final manuscript, and agree to be accountable for all aspects of the work.

---

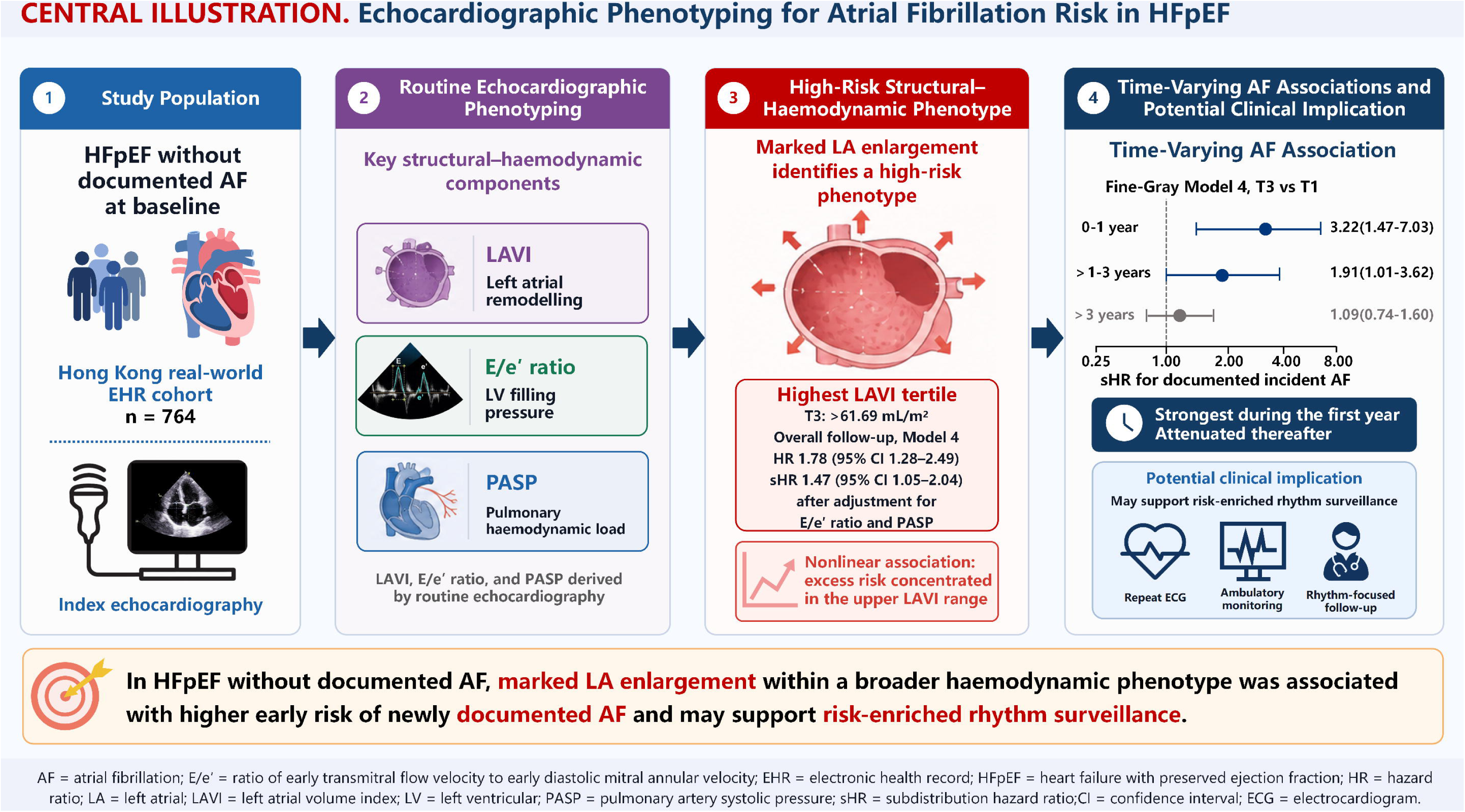

## Notes

### Competing Interest Statement

The authors have declared no competing interest.

### Author Declarations

The Joint Chinese University of Hong Kong-New Territories East Cluster Clinical Research Ethics Committee gave ethical approval for this work (approval number 2019.422). The Committee waived the requirement for individual informed consent because the study was retrospective and used deidentified electronic health record data without patient contact.

