## Supplementary methods, tables, and figures for "Left Atrial Remodeling, Hemodynamic Burden, and Time-Varying Risk of Newly Documented Atrial Fibrillation in Heart Failure With Preserved Ejection Fraction"

***Data preparation and variable definitions***

The first eligible echocardiographic examination during the study period was used to define the index date and baseline echocardiographic phenotype. Clinical covariates and medication use were ascertained from structured records available at or before the index examination. Diagnostic variables were defined using prespecified CDARS coding algorithms used for HFpEF cohort construction^1-5^.

Atrial fibrillation (AF)-related variables were defined from routinely documented inpatient and outpatient diagnoses in CDARS and available electrocardiographic records. Pre-existing AF referred to documented AF on or before the index echocardiographic examination, whereas incident AF referred to the first qualifying AF diagnosis or electrocardiographic record after the index date. Because systematic rhythm monitoring was not performed before or after the index examination, the incident AF endpoint in these analyses should be interpreted as clinically documented incident AF, or newly documented AF in routine care, rather than systematically detected de novo AF onset.

Values of zero for physiologically positive echocardiographic measurements were considered implausible and were recoded as missing. Continuous echocardiographic variables were retained on their original scales for the primary analyses. Left atrial volume index (LAVI) was rescaled to represent the effect per 10-mL/m² increase. For categorical analyses, fixed tertile cut points of 42.27 and 61.69 mL/m², derived from the observed pre-imputation LAVI distribution, were applied in all completed datasets and sensitivity analyses. Descriptive baseline summaries and cumulative incidence estimates according to LAVI tertiles were based on patients with observed pre-imputation LAVI measurements, whereas overall cumulative incidence estimates were calculated in the full analytic cohort.

For the echocardiographic phenotype analyses, LAVI, E/e′ ratio, and pulmonary artery systolic pressure (PASP) were standardised using their respective standard deviations calculated from the observed pre-imputation distributions. Each phenotype was evaluated in a separate model using the Model 3 covariate set.

***Multiple imputation***

The amount and pattern of missingness were summarised before imputation. Multiple imputation by chained equations was used under a missing-at-random assumption, and 30 imputed datasets were generated. The imputation model included all demographic, clinical, medication, and echocardiographic variables used in the primary or sensitivity analyses, together with competing-risk outcome status and follow-up time. Patient identifiers were not used as predictors, and outcome status and follow-up time were not imputed.

Type-appropriate conditional models were used for continuous and binary variables. Derived variables, including LAVI per 10 mL/m², LAVI tertiles, standardised echocardiographic phenotypes, and model-specific interaction terms, were reconstructed after completion of each imputed dataset rather than imputed directly.

All regression models were fitted independently within each of the 30 completed datasets. Regression coefficients and their covariance matrices were pooled according to Rubin’s rules. Imputation stability was assessed by examining convergence across imputation chains and comparing the distributions of observed and imputed values. A complete-case sensitivity analysis was conducted using a common cohort with complete data for LAVI and all covariates included in Model 4.

***Restricted cubic spline analyses***

Potential nonlinear associations between LAVI and documented incident AF were assessed using restricted cubic splines with 4 knots placed at the 5th, 35th, 65th, and 95th percentiles of the observed pre-imputation LAVI distribution. The median of the observed LAVI distribution was used as the reference value.

Spline terms were incorporated into cause-specific Cox and Fine–Gray models using the Model 3 and Model 4 covariate sets. Overall associations and departures from linearity were assessed using pooled joint Wald tests across the imputed datasets. For graphical presentation, estimated curves and 95% confidence intervals were displayed within the 1st to 99th percentiles of the observed LAVI distribution to reduce instability arising from sparse observations at the extremes.

The spline analyses were repeated after winsorising LAVI at the 1st and 99th percentiles and after excluding participants with observed LAVI values above the 99th percentile.

***Proportional hazards and time-varying analyses***

The proportional hazards assumption for the cause-specific Cox models was assessed using scaled Schoenfeld residuals. Formal tests were performed for the LAVI term and for the global model, and smoothed residual plots were used to examine the direction and timing of any change in the estimated association. These diagnostic analyses were performed in the common complete-case cohort to ensure that the same analytic sample was used across models.

To estimate period-specific associations, follow-up was divided at 1 and 3 years, generating intervals of 0–1 year, greater than 1–3 years, and greater than 3 years. Extended cause-specific Cox models were fitted after splitting each participant’s follow-up according to these intervals. The baseline hazard was stratified by follow-up period, and interactions between LAVI and period were used to estimate interval-specific HRs.

Time-varying Fine–Gray models were fitted over the same intervals using piecewise-constant subdistribution effects defined by step functions at 1 and 3 years. Death before documented incident AF remained the competing event. The period-specific Cox and Fine–Gray models were fitted in each imputed dataset, and coefficients and covariance matrices were pooled according to Rubin’s rules.

Time interaction was evaluated using pooled joint Wald tests. For continuous LAVI, the null hypothesis was equality of the 3 period-specific coefficients, corresponding to a 2-degree-of-freedom test. For LAVI tertiles, the interaction test jointly assessed the time-varying effects of T2 and T3 and had 4 degrees of freedom. Period-specific cause-specific Cox models were additionally repeated in the complete-case cohort.

***Landmark analyses***

Landmark analyses were conducted at 30 days, 90 days, and 1 year after the index echocardiographic examination. At each landmark, participants who had experienced documented incident AF, competing death, or censoring on or before the landmark were excluded. Eligible participants were required to remain alive, without documented incident AF, and under follow-up beyond the landmark date. Follow-up time was reset to zero at the corresponding landmark.

Within each landmark cohort, LAVI was evaluated both continuously and according to the prespecified tertiles using Model 3 and Model 4 cause-specific Cox and Fine–Gray regression. These analyses were undertaken to reduce the potential influence of previously unrecognised AF or very early post-index AF documentation, and to examine whether the association between LAVI and subsequently documented AF persisted beyond the early post-index period.

***Additional sensitivity and secondary analyses***

Complete-case analyses repeated the progressive Model 1 to Model 4 adjustment strategy using a fixed cohort with complete data for the primary exposure and all Model 4 covariates.

The influence of extreme LAVI values was examined using 2 complementary approaches. In the first, observed and imputed LAVI values were winsorised at the 1st and 99th percentiles of the observed pre-imputation distribution. In the second, participants with observed baseline LAVI above the 99th percentile were excluded from each completed dataset. Continuous Model 3 and Model 4 cause-specific Cox and Fine–Gray analyses were repeated under both approaches.

Left atrial diameter and the left atrial-to-aortic root ratio were evaluated as alternative measures of left atrial size. Each measure was standardised using its observed pre-imputation standard deviation and assessed in a separate Model 3 cause-specific Cox and Fine–Gray model.

A treatment-adjusted sensitivity model included all Model 4 covariates together with baseline beta-blocker and diuretic use. This model was intended to assess whether adjustment for commonly prescribed heart failure and rate-controlling therapies materially altered the association between LAVI and documented incident AF.

Separate univariable cause-specific Cox and Fine–Gray models were used to describe associations between selected demographic, clinical, and echocardiographic variables and documented incident AF. Continuous variables were reported per 1-standard-deviation increase, and binary variables using clinically relevant contrasts. These analyses were descriptive and were not used for covariate selection.

Correlations among LAVI, left atrial diameter, the left atrial-to-aortic root ratio, E/e′ ratio, PASP, and left ventricular mass index were assessed using Spearman correlation coefficients. Multicollinearity in the most extensively adjusted model was evaluated using variance inflation factors.

Secondary phenotype comparisons, univariable analyses, and sensitivity analyses were considered supportive and exploratory. No adjustment for multiplicity was applied to these analyses.

**Supplementary Methods**

**Supplementary Table 1.** Baseline Characteristics According to Competing-Risk Outcome Status

| **Characteristic** | **Overall (N=764)** | **Event-free/censored (n=132)** | **Death before AF (n=272)** | **Incident AF (n=360)** | ***P* value** |
| --- | --- | --- | --- | --- | --- |
| **Demographic and clinical characteristics** | | | | | |
| Age, years | 75.9 ± 11.3 | 70.0 ± 14.3 | 78.5 ± 9.9 | 76.0 ± 10.3 | <0.001 |
| Male sex, n (%) | 346 (45.3) | 64 (48.5) | 136 (50.0) | 146 (40.6) | 0.044 |
| Heart rate, beats/min | 78.9 ± 20.1 | 78.1 ± 24.6 | 76.9 ± 16.3 | 80.8 ± 20.8 | 0.027 |
| **Comorbidities** | | | | | |
| Diabetes mellitus | 246 (32.2) | 45 (34.1) | 78 (28.7) | 123 (34.2) | 0.301 |
| Renal disease | 99 (13.0) | 21 (15.9) | 34 (12.5) | 44 (12.2) | 0.537 |
| Hypertension | 421 (55.1) | 72 (54.5) | 146 (53.7) | 203 (56.4) | 0.786 |
| Chronic obstructive pulmonary disease | 124 (16.2) | 20 (15.2) | 40 (14.7) | 64 (17.8) | 0.546 |
| Ischaemic heart disease | 349 (45.7) | 61 (46.2) | 126 (46.3) | 162 (45.0) | 0.938 |
| **Baseline medications** | | | | | |
| Beta-blocker use | 397 (52.0) | 61 (46.2) | 143 (52.6) | 193 (53.6) | 0.336 |
| Diuretic use | 390 (51.0) | 70 (53.0) | 128 (47.1) | 192 (53.3) | 0.260 |
| **Echocardiographic characteristics** | | | | | |
| Left ventricular ejection fraction, % | 60.9 ± 7.9 | 62.0 ± 7.6 | 61.4 ± 8.0 | 60.2 ± 8.0 | 0.031 |
| Left ventricular mass index, g/m² | 102.50 (85.16–143.84) | 95.51 (80.13–129.38) | 103.89 (88.36–146.46) | 102.85 (84.12–145.47) | 0.012 |
| Left atrial volume index, mL/m² | 43.00 (40.99–67.81) | 43.09 (40.41–59.77) | 42.63 (39.16–63.55) | 54.48 (42.26–85.45) | <0.001 |
| Left atrial diameter, cm | 3.49 (3.33–3.78) | 3.37 (3.25–3.50) | 3.45 (3.31–3.61) | 3.65 (3.40–4.02) | <0.001 |
| Left atrial-to-aortic root ratio | 1.42 (1.28–1.60) | 1.36 (1.26–1.47) | 1.33 (1.26–1.47) | 1.52 (1.35–1.73) | <0.001 |
| E/e′ ratio | 18.91 (16.28–21.66) | 17.63 (15.40–19.74) | 18.57 (15.73–20.98) | 19.76 (17.25–22.81) | <0.001 |
| Pulmonary artery systolic pressure, mm Hg | 39.92 (34.62–49.09) | 35.80 (33.42–40.00) | 37.21 (34.00–45.00) | 45.00 (36.00–53.00) | <0.001 |

Values were calculated from observed pre-imputation data and are presented as mean ± standard deviation, median (interquartile range), or n (%), as appropriate. P values compare the three outcome groups and were calculated using Welch one-way analysis of variance, the Kruskal–Wallis test, or the Pearson χ² test or Fisher exact test, as appropriate. The event-free/censored group included patients who remained alive and free from documented incident AF at the end of follow-up. The death-before-AF group included patients who died before documented incident AF. Documented incident AF was defined as the first qualifying AF diagnosis or electrocardiographic record after the index echocardiogram among patients without documented AF at baseline. Percentages use the corresponding group size as the denominator. P values are descriptive and were not adjusted for multiple comparisons. AF, atrial fibrillation; E/e′, ratio of early transmitral flow velocity to early diastolic mitral annular velocity; HFpEF, heart failure with preserved ejection fraction; LV, left ventricular.

**Supplementary Table 2.** Missingness of Variables Included in the Documented Incident Atrial Fibrillation Analyses

| **Variable** | **Available n** | **Overall missing (N=764), n (%)** | **No incident AF (n=404), n (%)** | **Incident AF (n=360), n (%)** |
| --- | --- | --- | --- | --- |
| **Demographic and clinical characteristics** | | | | |
| Age, years | 764 | 0 (0.0) | 0 (0.0) | 0 (0.0) |
| Male sex | 764 | 0 (0.0) | 0 (0.0) | 0 (0.0) |
| Heart rate, beats/min | 764 | 0 (0.0) | 0 (0.0) | 0 (0.0) |
| **Comorbidities** | | | | |
| Diabetes mellitus | 764 | 0 (0.0) | 0 (0.0) | 0 (0.0) |
| Renal disease | 764 | 0 (0.0) | 0 (0.0) | 0 (0.0) |
| Hypertension | 764 | 0 (0.0) | 0 (0.0) | 0 (0.0) |
| Chronic obstructive pulmonary disease | 764 | 0 (0.0) | 0 (0.0) | 0 (0.0) |
| Ischaemic heart disease | 764 | 0 (0.0) | 0 (0.0) | 0 (0.0) |
| **Baseline medications** | | | | |
| Beta-blocker use | 764 | 0 (0.0) | 0 (0.0) | 0 (0.0) |
| Diuretic use | 764 | 0 (0.0) | 0 (0.0) | 0 (0.0) |
| **Echocardiographic characteristics** | | | | |
| Left ventricular ejection fraction, % | 761 | 3 (0.4) | 0 (0.0) | 3 (0.8) |
| Left ventricular mass index, g/m² | 716 | 48 (6.3) | 32 (7.9) | 16 (4.4) |
| Left atrial volume index, mL/m² | 715 | 49 (6.4) | 22 (5.4) | 27 (7.5) |
| Left atrial diameter, cm | 722 | 42 (5.5) | 20 (5.0) | 22 (6.1) |
| Left atrial-to-aortic root ratio | 703 | 61 (8.0) | 30 (7.4) | 31 (8.6) |
| E/e′ ratio | 735 | 29 (3.8) | 15 (3.7) | 14 (3.9) |
| Pulmonary artery systolic pressure, mm Hg | 720 | 44 (5.8) | 23 (5.7) | 21 (5.8) |

Missing values are presented as n (%). Percentages were calculated using the number of participants in the corresponding column as the denominator. Available n denotes the number of participants with nonmissing values before multiple imputation. The no documented incident AF group comprised participants who remained free from documented incident AF and those who died before documented incident AF. Variables shown were those included in the primary clinical model, the haemodynamic overlap-adjusted model, alternative echocardiographic phenotype analyses, or treatment-adjusted sensitivity analysis. For physiologically positive continuous echocardiographic measurements, values of zero were treated as missing. AF, atrial fibrillation; LV, left ventricular.

**Supplementary Table 3.** Cumulative Incidence of Documented Incident Atrial Fibrillation According to Left Atrial Volume Index Tertiles

| **Group** | **N** | **Documented incident AF, n** | **Death before documented incident AF, n** | **Event-free or censored, n** | **1-year cumulative incidence, % (95% CI)** | **3-year cumulative incidence, % (95% CI)** | **5-year cumulative incidence, % (95% CI)** |
| --- | --- | --- | --- | --- | --- | --- | --- |
| Overall | 764 | 360 | 272 | 132 | 7.3 (5.5–9.2) | 17.8 (15.1–20.6) | 26.7 (23.6–29.9) |
| T1 | 239 | 85 | 111 | 43 | 3.3 (1.1–5.6) | 8.8 (5.2–12.4) | 15.6 (11.0–20.3) |
| T2 | 238 | 104 | 80 | 54 | 3.8 (1.4–6.2) | 13.0 (8.8–17.3) | 19.9 (14.8–25.0) |
| T3 | 238 | 144 | 71 | 23 | 16.0 (11.3–20.6) | 30.7 (24.9–36.6) | 43.4 (37.1–49.8) |

Values are presented as n or cumulative incidence percentage (95% CI). Documented incident AF was defined as the first qualifying AF diagnosis or electrocardiographic record after the index echocardiogram among patients without documented AF at baseline. Death before documented incident AF was treated as a competing event. Overall estimates were calculated in the full analytic cohort of 764 patients, whereas LAVI-tertile estimates were calculated among 715 patients with observed pre-imputation LAVI measurements. LAVI tertiles were defined as T1, ≤42.27 mL/m²; T2, >42.27 to ≤61.69 mL/m²; and T3, >61.69 mL/m². Gray’s test for differences across LAVI tertiles was P<0.001. AF, atrial fibrillation; CI, confidence interval; LAVI, left atrial volume index.

**Supplementary Table 4.** Adjusted Associations of Selected Echocardiographic Phenotypes With Documented Incident Atrial Fibrillation

| **Phenotype** | **Domain** | **Observed n before imputation** | **Scaling or contrast** | **Cause-specific HR (95% CI)** | **Cause-specific *P* value** | **Fine–Gray sHR (95% CI)** | **Fine–Gray *P* value** |
| --- | --- | --- | --- | --- | --- | --- | --- |
| Left atrial volume index | Left atrial structure | 715 | Per 1 SD (35.82) increase | 1.33 (1.22–1.46) | <0.001 | 1.25 (1.13–1.37) | <0.001 |
| E/e′ ratio | LV filling pressure | 735 | Per 1 SD (6.72) increase | 1.26 (1.17–1.35) | <0.001 | 1.20 (1.13–1.28) | <0.001 |
| Pulmonary artery systolic pressure | Pulmonary haemodynamic load | 720 | Per 1 SD (10.87) increase | 1.51 (1.39–1.64) | <0.001 | 1.42 (1.26–1.60) | <0.001 |

Values are hazard ratios or subdistribution hazard ratios with 95% confidence intervals per 1-standard-deviation increase in the corresponding echocardiographic phenotype. Documented incident AF was defined as the first qualifying AF diagnosis or electrocardiographic record after the index echocardiogram among patients without documented AF at baseline. All models included 764 participants after multiple imputation; observed n before imputation denotes the number of participants with nonmissing values for the corresponding phenotype before imputation. Standard deviations were calculated from the observed pre-imputation distributions. Each echocardiographic phenotype was evaluated in a separate Model 3, the primary clinical adjustment model, which adjusted for age, sex, heart rate, left ventricular ejection fraction, left ventricular mass index, diabetes mellitus, renal disease, hypertension, chronic obstructive pulmonary disease, and ischaemic heart disease. Cause-specific Cox regression treated death before documented incident AF as censoring, whereas Fine–Gray regression treated death before documented incident AF as a competing event. Missing covariate data were handled using multiple imputation by chained equations with 30 imputed data sets, and estimates were pooled according to Rubin’s rules. P values were not adjusted for multiple comparisons. AF, atrial fibrillation; CI, confidence interval; COPD, chronic obstructive pulmonary disease; HR, hazard ratio; IHD, ischaemic heart disease; LV, left ventricular; LVEF, left ventricular ejection fraction; SD, standard deviation; sHR, subdistribution hazard ratio.

**Supplementary Table 5.** Complete-Case Sensitivity Analysis of the Association Between Left Atrial Volume Index and Documented Incident Atrial Fibrillation

| **Exposure** | **Model** | **Cause-specific HR (95% CI)** | ***P* value** | **Fine–Gray sHR (95% CI)** | ***P* value** |
| --- | --- | --- | --- | --- | --- |
| **Panel A. Left atrial volume index as a continuous variable** | | | | | |
| Per 10 mL/m² increase | Model 1 | 1.08 (1.05–1.11) | <0.001 | 1.07 (1.04–1.10) | <0.001 |
| Per 10 mL/m² increase | Model 2 | 1.08 (1.05–1.11) | <0.001 | 1.07 (1.04–1.10) | <0.001 |
| Per 10 mL/m² increase | Model 3 (primary clinical) | 1.08 (1.05–1.11) | <0.001 | 1.06 (1.03–1.10) | <0.001 |
| Per 10 mL/m² increase | Model 4 (haemodynamic overlap-adjusted) | 1.02 (0.99–1.06) | 0.195 | 1.02 (0.98–1.05) | 0.410 |
| **Panel B. Left atrial volume index tertiles** | | | | | |
| T1 | Model 3 (primary clinical) | 1.00 (Reference) | — | 1.00 (Reference) | — |
| T2 | Model 3 (primary clinical) | 1.16 (0.84–1.61) | 0.360 | 1.27 (0.93–1.75) | 0.138 |
| T3 | Model 3 (primary clinical) | 2.73 (1.98–3.78) | <0.001 | 2.28 (1.66–3.12) | <0.001 |
| Overall P | Model 3 (primary clinical) | — | <0.001 | — | <0.001 |
| P for trend | Model 3 (primary clinical) | — | <0.001 | — | <0.001 |
| T1 | Model 4 (haemodynamic overlap-adjusted) | 1.00 (Reference) | — | 1.00 (Reference) | — |
| T2 | Model 4 (haemodynamic overlap-adjusted) | 0.98 (0.70–1.37) | 0.911 | 1.12 (0.81–1.55) | 0.505 |
| T3 | Model 4 (haemodynamic overlap-adjusted) | 1.79 (1.25–2.56) | 0.001 | 1.59 (1.12–2.27) | 0.010 |
| Overall P | Model 4 (haemodynamic overlap-adjusted) | — | <0.001 | — | 0.018 |
| P for trend | Model 4 (haemodynamic overlap-adjusted) | — | <0.001 | — | 0.008 |

Sensitivity analyses were restricted to patients with complete data for LAVI and all variables included in Model 4. The same fixed complete-case cohort was used for all progressive models. The cohort comprised 609 patients, including 287 documented incident AF events and 217 deaths before documented incident AF. Documented incident AF was defined as the first qualifying AF diagnosis or electrocardiographic record after the index echocardiogram among patients without documented AF at baseline. Cause-specific Cox regression treated death before documented incident AF as censoring, whereas Fine–Gray regression treated death before documented incident AF as a competing event. LAVI tertiles were defined using the fixed cut points of 42.27 and 61.69 mL/m² derived from the observed pre-imputation distribution; T1 was the reference category. Overall P values were obtained from joint 2-degree-of-freedom Wald tests of the T2 and T3 coefficients, and P for trend was obtained by modelling tertile rank as an ordinal variable.

Model 1 was adjusted for age and sex. Model 2 was additionally adjusted for heart rate. Model 3, the primary clinical model, additionally included left ventricular ejection fraction, left ventricular mass index, diabetes mellitus, renal disease, hypertension, chronic obstructive pulmonary disease, and ischaemic heart disease. Model 4, the haemodynamic overlap-adjusted model, additionally included E/e′ ratio and pulmonary artery systolic pressure. AF, atrial fibrillation; CI, confidence interval; HR, hazard ratio; LAVI, left atrial volume index; sHR, subdistribution hazard ratio.

**Supplementary Table 6.** Landmark Analyses at 30 Days, 90 Days, and 1 Year of the Association Between Left Atrial Volume Index and Documented Incident Atrial Fibrillation

| **Analysis** | **Exposure** | **Model** | **Cause-specific HR (95% CI)** | ***P* value** | **Fine-Gray sHR (95% CI)** | ***P* value** |
| --- | --- | --- | --- | --- | --- | --- |
| **30-day landmark (eligible n = 731)** | | | | | | |
| Continuous LAVI | Per 10 mL/m² increase | Model 3 (primary clinical) | 1.08 (1.05–1.11) | <0.001 | 1.06 (1.03–1.09) | <0.001 |
|  | Per 10 mL/m² increase | Model 4 (haemodynamic overlap-adjusted) | 1.02 (0.99–1.05) | 0.302 | 1.02 (0.98–1.05) | 0.343 |
| LAVI tertiles | T1 | Model 3 (primary clinical) | 1.00 (Reference) | — | 1.00 (Reference) | — |
|  | T2 |  | 1.21 (0.90–1.62) | 0.210 | 1.28 (0.96–1.71) | 0.096 |
|  | T3 |  | 2.59 (1.91–3.50) | <0.001 | 2.27 (1.69–3.06) | <0.001 |
|  | Overall P |  | — | <0.001 | — | <0.001 |
|  | P for trend |  | — | <0.001 | — | <0.001 |
|  | T1 | Model 4 (haemodynamic overlap-adjusted) | 1.00 (Reference) | — | 1.00 (Reference) | — |
|  | T2 |  | 1.02 (0.75–1.37) | 0.915 | 1.12 (0.83–1.51) | 0.461 |
|  | T3 |  | 1.62 (1.15–2.28) | 0.006 | 1.56 (1.11–2.20) | 0.011 |
|  | Overall P |  | — | 0.003 | — | 0.024 |
|  | P for trend |  | — | 0.005 | — | 0.010 |
| **90-day landmark (eligible n = 716)** | | | | | | |
| Continuous LAVI | Per 10 mL/m² increase | Model 3 (primary clinical) | 1.07 (1.04–1.10) | <0.001 | 1.06 (1.03–1.09) | <0.001 |
|  | Per 10 mL/m² increase | Model 4 (haemodynamic overlap-adjusted) | 1.00 (0.97–1.04) | 0.803 | 1.01 (0.97–1.04) | 0.613 |
| LAVI tertiles | T1 | Model 3 (primary clinical) | 1.00 (Reference) | — | 1.00 (Reference) | — |
|  | T2 |  | 1.20 (0.90–1.62) | 0.219 | 1.28 (0.96–1.71) | 0.093 |
|  | T3 |  | 2.50 (1.84–3.39) | <0.001 | 2.36 (1.75–3.19) | <0.001 |
|  | Overall P |  | — | <0.001 | — | <0.001 |
|  | P for trend |  | — | <0.001 | — | <0.001 |
|  | T1 | Model 4 (haemodynamic overlap-adjusted) | 1.00 (Reference) | — | 1.00 (Reference) | — |
|  | T2 |  | 1.01 (0.75–1.37) | 0.925 | 1.12 (0.83–1.51) | 0.442 |
|  | T3 |  | 1.57 (1.11–2.22) | 0.010 | 1.64 (1.16–2.31) | 0.005 |
|  | Overall P |  | — | 0.007 | — | 0.011 |
|  | P for trend |  | — | 0.009 | — | 0.005 |
| **1-year landmark (eligible n = 642)** | | | | | | |
| Continuous LAVI | Per 10 mL/m² increase | Model 3 (primary clinical) | 1.06 (1.03–1.09) | <0.001 | 1.06 (1.03–1.09) | <0.001 |
|  | Per 10 mL/m² increase | Model 4 (haemodynamic overlap-adjusted) | 1.00 (0.96–1.03) | 0.843 | 1.01 (0.97–1.05) | 0.580 |
| LAVI tertiles | T1 | Model 3 (primary clinical) | 1.00 (Reference) | — | 1.00 (Reference) | — |
|  | T2 |  | 1.16 (0.85–1.60) | 0.345 | 1.35 (0.98–1.85) | 0.066 |
|  | T3 |  | 2.36 (1.70–3.28) | <0.001 | 2.64 (1.90–3.66) | <0.001 |
|  | Overall P |  | — | <0.001 | — | <0.001 |
|  | P for trend |  | — | <0.001 | — | <0.001 |
|  | T1 | Model 4 (haemodynamic overlap-adjusted) | 1.00 (Reference) | — | 1.00 (Reference) | — |
|  | T2 |  | 0.98 (0.71–1.35) | 0.917 | 1.19 (0.86–1.64) | 0.298 |
|  | T3 |  | 1.50 (1.03–2.17) | 0.034 | 1.92 (1.32–2.78) | <0.001 |
|  | Overall P |  | — | 0.022 | — | 0.001 |
|  | P for trend |  | — | 0.032 | — | <0.001 |

Landmark analyses included patients who remained alive, free from documented incident AF, and under follow-up beyond the corresponding landmark. Patients whose documented incident AF, competing death, or censoring occurred on or before the corresponding landmark were excluded, and follow-up time was reset to zero at the landmark. The 30-day cohort included 731 patients, 345 subsequent documented incident AF events, and 254 deaths before documented incident AF; the 90-day cohort included 716 patients, 337 subsequent documented incident AF events, and 247 deaths before documented incident AF; and the 1-year cohort included 642 patients, 304 subsequent documented incident AF events, and 206 deaths before documented incident AF. Model 3, the primary clinical model, adjusted for age, sex, heart rate, left ventricular ejection fraction, left ventricular mass index, diabetes mellitus, renal disease, hypertension, chronic obstructive pulmonary disease, and ischaemic heart disease. Model 4, the haemodynamic overlap-adjusted model, additionally adjusted for E/e′ ratio and pulmonary artery systolic pressure. Cause-specific Cox regression treated death before documented incident AF as censoring, whereas Fine–Gray regression treated death before documented incident AF as a competing event. Missing baseline covariates were handled using multiple imputation by chained equations with 30 imputed data sets, and estimates were pooled using Rubin’s rules. LAVI tertiles were defined using fixed pre-imputation cut points of 42.27 and 61.69 mL/m²; T1 was the reference category. Overall P values were obtained using joint 2-degree-of-freedom Wald tests of the T2 and T3 coefficients, and P for trend was obtained by modelling tertile rank as an ordinal variable. AF, atrial fibrillation; CI, confidence interval; COPD, chronic obstructive pulmonary disease; HR, hazard ratio; IHD, ischaemic heart disease; LAVI, left atrial volume index; LVEF, left ventricular ejection fraction; PASP, pulmonary artery systolic pressure; sHR, subdistribution hazard ratio.

**Supplementary Table 7.** Sensitivity Analyses Using Alternative Measures of Left Atrial Size and Documented Incident Atrial Fibrillation

| **Measure** | **Observed n before imputation** | **Scaling or contrast** | **Cause-specific HR (95% CI)** | ***P* value** | **Fine–Gray sHR (95% CI)** | ***P* value** |
| --- | --- | --- | --- | --- | --- | --- |
| Left atrial diameter | 722 | Per 1-SD (0.45 cm) increase | 1.40 (1.28–1.53) | <0.001 | 1.46 (1.29–1.64) | <0.001 |
| Left atrial-to-aortic root ratio | 703 | Per 1-SD (0.27) increase | 1.40 (1.27–1.54) | <0.001 | 1.53 (1.38–1.69) | <0.001 |

Values are hazard ratios or subdistribution hazard ratios with 95% confidence intervals per 1-standard-deviation increase in the corresponding left atrial measure. Documented incident AF was defined as the first qualifying AF diagnosis or electrocardiographic record after the index echocardiogram among patients without documented AF at baseline. Standard deviations were calculated from the observed pre-imputation distributions. All models included 764 participants after multiple imputation; observed n before imputation represents the number of participants with nonmissing values for the corresponding measure before imputation. Each alternative left atrial measure was evaluated in a separate Model 3, the primary clinical model, adjusted for age, sex, heart rate, left ventricular ejection fraction, left ventricular mass index, diabetes mellitus, renal disease, hypertension, chronic obstructive pulmonary disease, and ischaemic heart disease. Cause-specific Cox regression treated death before documented incident AF as censoring, whereas Fine–Gray regression treated death before documented incident AF as a competing event. Missing covariate data were handled using multiple imputation by chained equations with 30 imputed data sets, and estimates were pooled using Rubin’s rules. P values were not adjusted for multiple comparisons. AF, atrial fibrillation; CI, confidence interval; HR, hazard ratio; LA/Ao, left atrial-to-aortic root ratio; LAD, left atrial diameter; LVEF, left ventricular ejection fraction; SD, standard deviation; sHR, subdistribution hazard ratio.

**Supplementary Table 8.** Extreme-Value Sensitivity Analyses of the Association Between Left Atrial Volume Index and Documented Incident Atrial Fibrillation

| **Method** | **Model** | **Eligible n** | **Exposure** | **Cause-specific HR (95% CI)** | ***P* value** | **Fine–Gray sHR (95% CI)** | ***P* value** |
| --- | --- | --- | --- | --- | --- | --- | --- |
| Winsorized at the 1st and 99th percentiles | Model 3 (primary clinical) | 764 | Per 10 mL/m² increase | 1.08 (1.06–1.11) | <0.001 | 1.06 (1.04–1.09) | <0.001 |
|  | Model 4 (haemodynamic overlap-adjusted) | 764 | Per 10 mL/m² increase | 1.03 (1.00–1.06) | 0.094 | 1.02 (0.98–1.05) | 0.359 |
| Excluded observed LAVI values above the 99th percentile | Model 3 (primary clinical) | 757 | Per 10 mL/m² increase | 1.10 (1.07–1.14) | <0.001 | 1.07 (1.04–1.10) | <0.001 |
|  | Model 4 (haemodynamic overlap-adjusted) | 757 | Per 10 mL/m² increase | 1.04 (1.01–1.08) | 0.011 | 1.02 (0.99–1.06) | 0.182 |

Values are hazard ratios or subdistribution hazard ratios with 95% confidence intervals per 10-mL/m² increase in left atrial volume index. Documented incident AF was defined as the first qualifying AF diagnosis or electrocardiographic record after the index echocardiogram among patients without documented AF at baseline. Two extreme-value strategies were examined. In the winsorized analyses, LAVI values were capped at the 1st and 99th percentiles of the observed pre-imputation distribution. In the exclusion analyses, participants with observed pre-imputation LAVI values above the 99th percentile were excluded from every imputed data set. Model 3, the primary clinical model, adjusted for age, sex, heart rate, left ventricular ejection fraction, left ventricular mass index, diabetes mellitus, renal disease, hypertension, chronic obstructive pulmonary disease, and ischaemic heart disease. Model 4, the haemodynamic overlap-adjusted model, additionally adjusted for E/e′ ratio and pulmonary artery systolic pressure. Cause-specific Cox regression treated death before documented incident AF as censoring, whereas Fine–Gray regression treated death before documented incident AF as a competing event. Missing covariate data were handled using multiple imputation by chained equations with 30 imputed data sets, and estimates were pooled using Rubin’s rules. AF, atrial fibrillation; CI, confidence interval; COPD, chronic obstructive pulmonary disease; HR, hazard ratio; IHD, ischaemic heart disease; LAVI, left atrial volume index; LVEF, left ventricular ejection fraction; PASP, pulmonary artery systolic pressure; sHR, subdistribution hazard ratio.

**Supplementary Table 9.** Treatment-Adjusted Sensitivity Analysis of the Association Between Left Atrial Volume Index and Documented Incident Atrial Fibrillation

| **Exposure** | **Model** | **Cause-specific HR (95% CI)** | ***P* value** | **Fine-Gray sHR (95% CI)** | ***P* value** |
| --- | --- | --- | --- | --- | --- |
| **Panel A. Left atrial volume index as a continuous variable** | | | | | |
| Per 10 mL/m² increase | Model 5 (treatment-adjusted sensitivity) | 1.03 (1.00-1.06) | 0.096 | 1.02 (0.98-1.05) | 0.384 |
| **Panel B. Left atrial volume index tertiles** | | | | | |
| T1 | Model 5 (treatment-adjusted sensitivity) | 1.00 (Reference) | — | 1.00 (Reference) | — |
| T2 | Model 5 (treatment-adjusted sensitivity) | 1.02 (0.76-1.38) | 0.877 | 1.08 (0.80-1.45) | 0.627 |
| T3 | Model 5 (treatment-adjusted sensitivity) | 1.79 (1.28-2.49) | <0.001 | 1.46 (1.05-2.03) | 0.025 |
| Overall P | Model 5 (treatment-adjusted sensitivity) | — | <0.001 | — | 0.046 |
| P for trend | Model 5 (treatment-adjusted sensitivity) | — | <0.001 | — | 0.022 |

Analyses included 764 patients, including 360 documented incident AF events and 272 deaths before documented incident AF. Documented incident AF was defined as the first qualifying AF diagnosis or electrocardiographic record after the index echocardiogram among patients without documented AF at baseline. Model 5 included all variables in Model 4 and additionally adjusted for baseline beta-blocker and diuretic use. Model 4, the haemodynamic overlap-adjusted model, adjusted for age, sex, heart rate, left ventricular ejection fraction, left ventricular mass index, diabetes mellitus, renal disease, hypertension, chronic obstructive pulmonary disease, ischaemic heart disease, E/e′ ratio, and pulmonary artery systolic pressure. Cause-specific Cox regression treated death before documented incident AF as censoring, whereas Fine–Gray regression treated death before documented incident AF as a competing event. Missing baseline covariates were handled using multiple imputation by chained equations with 30 imputed data sets, and estimates were pooled using Rubin’s rules. LAVI tertiles were defined using the fixed pre-imputation cut points of 42.27 and 61.69 mL/m², with T1 as the reference category. Overall P values were obtained from joint 2-degree-of-freedom Wald tests of the T2 and T3 coefficients; P for trend was obtained by modelling tertile rank as an ordinal variable. AF, atrial fibrillation; CI, confidence interval; COPD, chronic obstructive pulmonary disease; HR, hazard ratio; IHD, ischaemic heart disease; LAVI, left atrial volume index; LVEF, left ventricular ejection fraction; PASP, pulmonary artery systolic pressure; sHR, subdistribution hazard ratio.

**Supplementary Table 10.** Assessment of the Proportional Hazards Assumption for Cause-Specific Cox Regression Models Using Scaled Schoenfeld Residuals

| **Model** | **LAVI specification** | **Test** | **χ²** | **Degrees of freedom** | ***P* value** |
| --- | --- | --- | --- | --- | --- |
| Model 3 | Continuous LAVI, per 10 mL/m² | LAVI term | 4.61 | 1 | 0.032 |
|  |  | Global model test | 13.42 | 11 | 0.267 |
|  | LAVI tertiles | T2 vs T1 | 6.44 | 1 | 0.011 |
|  |  | T3 vs T1 | 11.61 | 1 | <0.001 |
|  |  | Overall tertile effect | 11.99 | 2 | 0.002 |
|  |  | Global model test | 21.55 | 12 | 0.043 |
| Model 4 | Continuous LAVI, per 10 mL/m² | LAVI term | 6.45 | 1 | 0.011 |
|  |  | Global model test | 17.58 | 13 | 0.174 |
|  | LAVI tertiles | T2 vs T1 | 7.30 | 1 | 0.007 |
|  |  | T3 vs T1 | 11.38 | 1 | <0.001 |
|  |  | Overall tertile effect | 12.12 | 2 | 0.002 |
|  |  | Global model test | 23.36 | 14 | 0.055 |

The proportional hazards assumption was assessed in the common complete-case sample (n=609; documented incident AF events, n=287). Individual tertile contrasts were evaluated at the coefficient level, whereas the overall tertile effect was evaluated using a 2-degree-of-freedom term-level test. Model 3, the primary clinical model, adjusted for age, sex, heart rate, left ventricular ejection fraction, left ventricular mass index, diabetes mellitus, renal disease, hypertension, chronic obstructive pulmonary disease, and ischaemic heart disease. Model 4, the haemodynamic overlap-adjusted model, additionally adjusted for E/e′ ratio and pulmonary artery systolic pressure. A P value <0.05 indicates evidence of potential non-proportionality. AF, atrial fibrillation; LAVI, left atrial volume index.

**Supplementary Table 11.** Period-Specific Associations of Left Atrial Volume Index With Documented Incident Atrial Fibrillation in Cause-Specific Cox Regression Models

| **Model** | **LAVI specification** | **Comparison** | **Follow-up period** | **Patients entering interval, n** | **Incident AF events, n** | **HR (95% CI)** | ***P* value** | ***P* for time interaction** |
| --- | --- | --- | --- | --- | --- | --- | --- | --- |
| Model 3 | Continuous LAVI | Per 10 mL/m² increase | 0–1 year | 609 | 47 | 1.16 (1.10–1.22) | <0.001 | 0.017 |
|  |  |  | >1–3 years | 509 | 65 | 1.08 (1.02–1.14) | 0.007 |  |
|  |  |  | >3 years | 388 | 175 | 1.05 (1.01–1.09) | 0.014 |  |
| Model 4 |  |  | 0–1 year | 609 | 47 | 1.10 (1.04–1.17) | <0.001 | 0.007 |
|  |  |  | >1–3 years | 509 | 65 | 1.02 (0.97–1.09) | 0.406 |  |
|  |  |  | >3 years | 388 | 175 | 0.99 (0.95–1.03) | 0.618 |  |
| Model 3 | LAVI tertiles | T2 vs T1 | 0–1 year | 609 | 47 | 0.85 (0.28–2.53) | 0.770 | 0.006 |
|  |  |  | >1–3 years | 509 | 65 | 1.57 (0.77–3.20) | 0.215 |  |
|  |  |  | >3 years | 388 | 175 | 1.08 (0.74–1.59) | 0.684 |  |
|  |  | T3 vs T1 | 0–1 year | 609 | 47 | 5.95 (2.61–13.55) | <0.001 |  |
|  |  |  | >1–3 years | 509 | 65 | 3.75 (1.91–7.36) | <0.001 |  |
|  |  |  | >3 years | 388 | 175 | 1.92 (1.28–2.88) | 0.002 |  |
| Model 4 |  | T2 vs T1 | 0–1 year | 609 | 47 | 0.71 (0.24–2.12) | 0.537 | 0.008 |
|  |  |  | >1–3 years | 509 | 65 | 1.31 (0.64–2.67) | 0.464 |  |
|  |  |  | >3 years | 388 | 175 | 0.92 (0.62–1.35) | 0.657 |  |
|  |  | T3 vs T1 | 0–1 year | 609 | 47 | 3.75 (1.63–8.66) | 0.002 |  |
|  |  |  | >1–3 years | 509 | 65 | 2.43 (1.22–4.84) | 0.011 |  |
|  |  |  | >3 years | 388 | 175 | 1.26 (0.81–1.95) | 0.306 |  |

Period-specific hazard ratios were estimated using extended cause-specific Cox regression models after splitting follow-up at 1 and 3 years, with the baseline hazard stratified by follow-up period. Death before documented incident AF was treated as a censoring event. The P value for time interaction was obtained using a likelihood ratio test comparing the period-specific model with the corresponding constant-effect model. For continuous LAVI, the interaction test had 2 degrees of freedom; for LAVI tertiles, the interaction test jointly evaluated the time-varying effects of T2 and T3 using 4 degrees of freedom. Analyses were performed in the common complete-case sample (n=609; documented incident AF events, n=287). Model 3, the primary clinical model, adjusted for age, sex, heart rate, left ventricular ejection fraction, left ventricular mass index, diabetes mellitus, renal disease, hypertension, chronic obstructive pulmonary disease, and ischaemic heart disease. Model 4, the haemodynamic overlap-adjusted model, additionally adjusted for E/e′ ratio and pulmonary artery systolic pressure. LAVI tertiles were defined as T1, ≤42.27 mL/m²; T2, >42.27 to ≤61.69 mL/m²; and T3, >61.69 mL/m². Patients at risk represent patients entering each follow-up interval. AF, atrial fibrillation; CI, confidence interval; HR, hazard ratio; LAVI, left atrial volume index.

**Supplementary Table 12.** Detailed Period-Specific Analyses of Left Atrial Volume Index and Documented Incident Atrial Fibrillation

| **Model** | **LAVI specification** | | **Comparison** | | **Follow-up period** | **Patients entering interval, n** | **Incident AF events, n** | **Cause-specific Cox regression** | | | **Fine–Gray regression** | | |
| --- | --- | --- | --- | --- | --- | --- | --- | --- | --- | --- | --- | --- | --- |
|  |  |  |  | |  | |  | **HR (95% CI)** | ***P* value** | ***P* for time interaction** | **sHR (95% CI)** | ***P* value** | **P for time interaction** |
| Model 3 | Continuous LAVI | Per 10 mL/m² increase | | 0–1 year | | 764 | 56 | 1.15 (1.10–1.20) | <0.001 | 0.004 | 1.14 (1.09–1.19) | <0.001 | 0.002 |
|  |  |  |  | >1–3 years | | 642 | 80 | 1.08 (1.03–1.13) | 0.002 |  | 1.05 (1.00–1.11) | 0.043 |  |
|  |  |  |  | >3 years | | 489 | 224 | 1.06 (1.02–1.10) | 0.002 |  | 1.04 (1.00–1.07) | 0.049 |  |
| Model 4 |  |  |  | 0–1 year | | 764 | 56 | 1.10 (1.05–1.15) | <0.001 | <0.001 | 1.09 (1.04–1.15) | <0.001 | 0.002 |
|  |  |  |  | >1–3 years | | 642 | 80 | 1.02 (0.97–1.07) | 0.438 |  | 1.00 (0.95–1.06) | 0.867 |  |
|  |  |  |  | >3 years | | 489 | 224 | 0.99 (0.96–1.03) | 0.779 |  | 0.99 (0.95–1.03) | 0.599 |  |
| Model 3 | LAVI tertiles | T2 vs T1 | | 0–1 year | | 764 | 56 | 1.20 (0.47–3.08) | 0.707 | 0.017 | 1.10 (0.43–2.84) | 0.841 | 0.007 |
|  |  |  |  | >1–3 years | | 642 | 80 | 1.67 (0.85–3.27) | 0.134 |  | 1.51 (0.78–2.93) | 0.220 |  |
|  |  |  |  | >3 years | | 489 | 224 | 1.06 (0.74–1.50) | 0.764 |  | 1.16 (0.82–1.63) | 0.396 |  |
|  |  | T3 vs T1 | | 0–1 year | | 764 | 56 | 6.09 (2.86–12.96) | <0.001 |  | 4.85 (2.26–10.37) | <0.001 |  |
|  |  |  |  | >1–3 years | | 642 | 80 | 3.80 (2.04–7.08) | <0.001 |  | 2.81 (1.53–5.18) | <0.001 |  |
|  |  |  |  | >3 years | | 489 | 224 | 2.03 (1.40–2.94) | <0.001 |  | 1.54 (1.08–2.19) | 0.017 |  |
| Model 4 |  | T2 vs T1 | | 0–1 year | | 764 | 56 | 0.99 (0.38–2.54) | 0.981 | 0.025 | 0.94 (0.36–2.42) | 0.898 | 0.011 |
|  |  |  |  | >1–3 years | | 642 | 80 | 1.36 (0.70–2.68) | 0.365 |  | 1.30 (0.67–2.53) | 0.436 |  |
|  |  |  |  | >3 years | | 489 | 224 | 0.90 (0.63–1.29) | 0.561 |  | 1.03 (0.72–1.46) | 0.885 |  |
|  |  | T3 vs T1 | | 0–1 year | | 764 | 56 | 3.77 (1.73–8.19) | <0.001 |  | 3.22 (1.47–7.03) | 0.003 |  |
|  |  |  |  | >1–3 years | | 642 | 80 | 2.38 (1.25–4.51) | 0.008 |  | 1.91 (1.01–3.62) | 0.047 |  |
|  |  |  |  | >3 years | | 489 | 224 | 1.31 (0.87–1.98) | 0.196 |  | 1.09 (0.74–1.60) | 0.666 |  |

Analyses included 764 patients, 360 documented incident AF events, and 272 deaths before documented incident AF. Patients entering each interval and documented incident AF events were obtained after splitting follow-up at 1 and 3 years. Cause-specific Cox regression treated death before documented incident AF as censoring, whereas Fine–Gray regression treated death before documented incident AF as a competing event. Estimates were obtained in 30 multiply imputed data sets and pooled using Rubin’s rules. Model 3, the primary clinical model, adjusted for age, sex, heart rate, left ventricular ejection fraction, left ventricular mass index, diabetes mellitus, renal disease, hypertension, chronic obstructive pulmonary disease, and ischaemic heart disease. Model 4, the haemodynamic overlap-adjusted model, additionally adjusted for E/e′ ratio and pulmonary artery systolic pressure. P values for time interaction were obtained from pooled joint Wald tests; continuous LAVI tests had 2 degrees of freedom and LAVI tertile tests had 4 degrees of freedom. LAVI tertiles were defined as T1, ≤42.27 mL/m²; T2, >42.27 to ≤61.69 mL/m²; and T3, >61.69 mL/m². AF, atrial fibrillation; CI, confidence interval; COPD, chronic obstructive pulmonary disease; HR, hazard ratio; IHD, ischaemic heart disease; LAVI, left atrial volume index; LVEF, left ventricular ejection fraction; PASP, pulmonary artery systolic pressure; sHR, subdistribution hazard ratio.

**Supplementary Figure 1.** Restricted Cubic Spline Associations of Left Atrial Volume Index With Documented Incident Atrial Fibrillation in the Haemodynamic Overlap-Adjusted Model

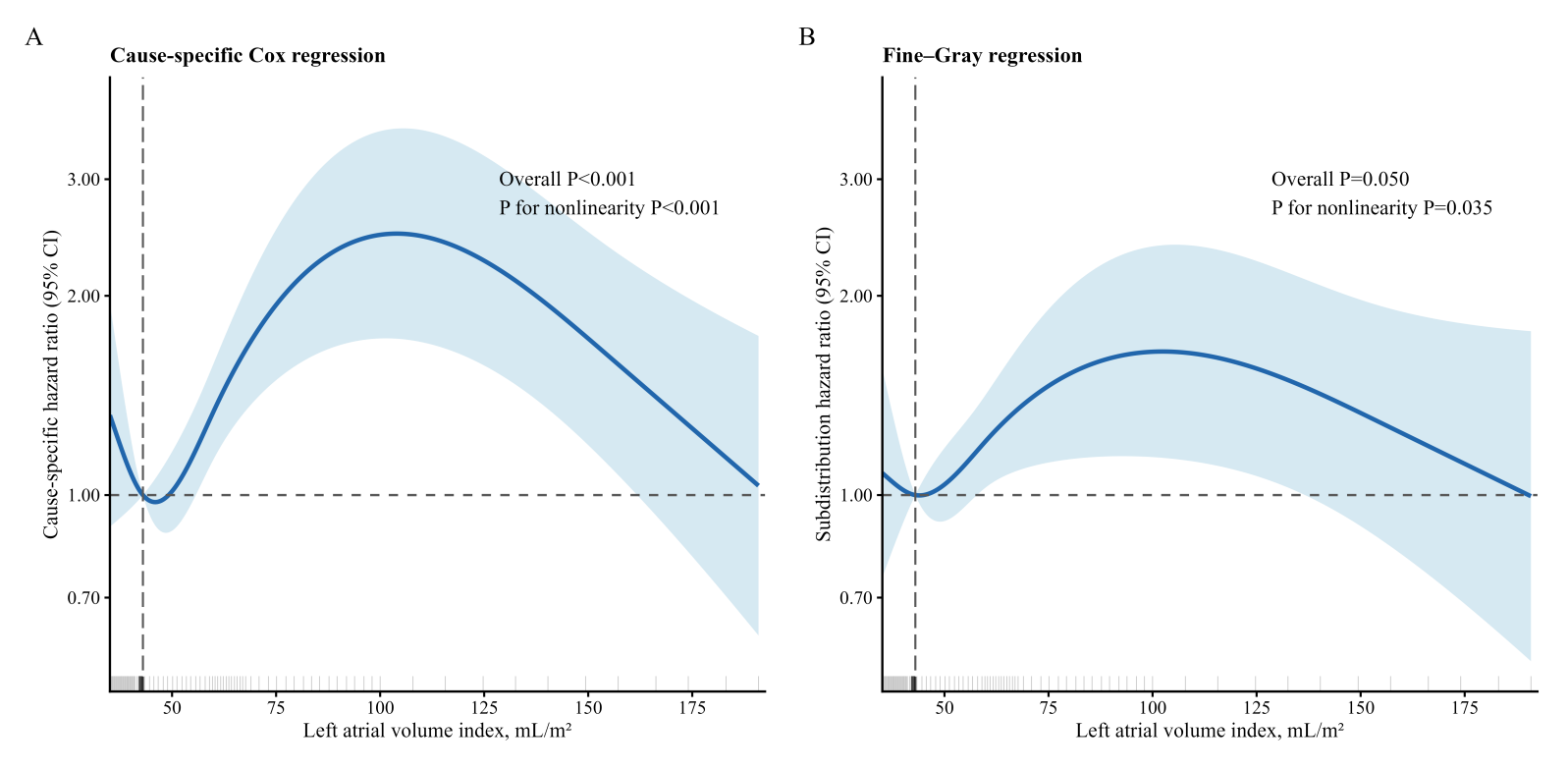

Restricted cubic spline curves depict the associations of left atrial volume index with documented incident AF using cause-specific Cox regression (A) and Fine–Gray competing-risk regression (B). Analyses were based on Model 4, the haemodynamic overlap-adjusted model, which adjusted for age, sex, heart rate, left ventricular ejection fraction, left ventricular mass index, diabetes mellitus, renal disease, hypertension, chronic obstructive pulmonary disease, ischaemic heart disease, E/e′ ratio, and pulmonary artery systolic pressure. Solid lines indicate the estimated hazard ratios or subdistribution hazard ratios, and shaded areas indicate the corresponding 95% confidence intervals. The horizontal dashed line denotes an effect estimate of 1.00, and the vertical dashed line indicates the reference LAVI value of 43.0 mL/m². Rug marks along the x-axis show the distribution of observed LAVI values. Overall and nonlinearity P values are displayed within each panel. Death before documented incident AF was treated as a censoring event in the cause-specific Cox model and as a competing event in the Fine–Gray model. Missing covariate data were handled using multiple imputation by chained equations with 30 imputed data sets, and estimates were pooled using Rubin’s rules. AF, atrial fibrillation; CI, confidence interval; E/e′, ratio of early transmitral flow velocity to early diastolic mitral annular velocity; LAVI, left atrial volume index; PASP, pulmonary artery systolic pressure.

**Supplementary Figure 2.** Extreme-Value Sensitivity Analyses of the Nonlinear Association Between Left Atrial Volume Index and Documented Incident Atrial Fibrillation

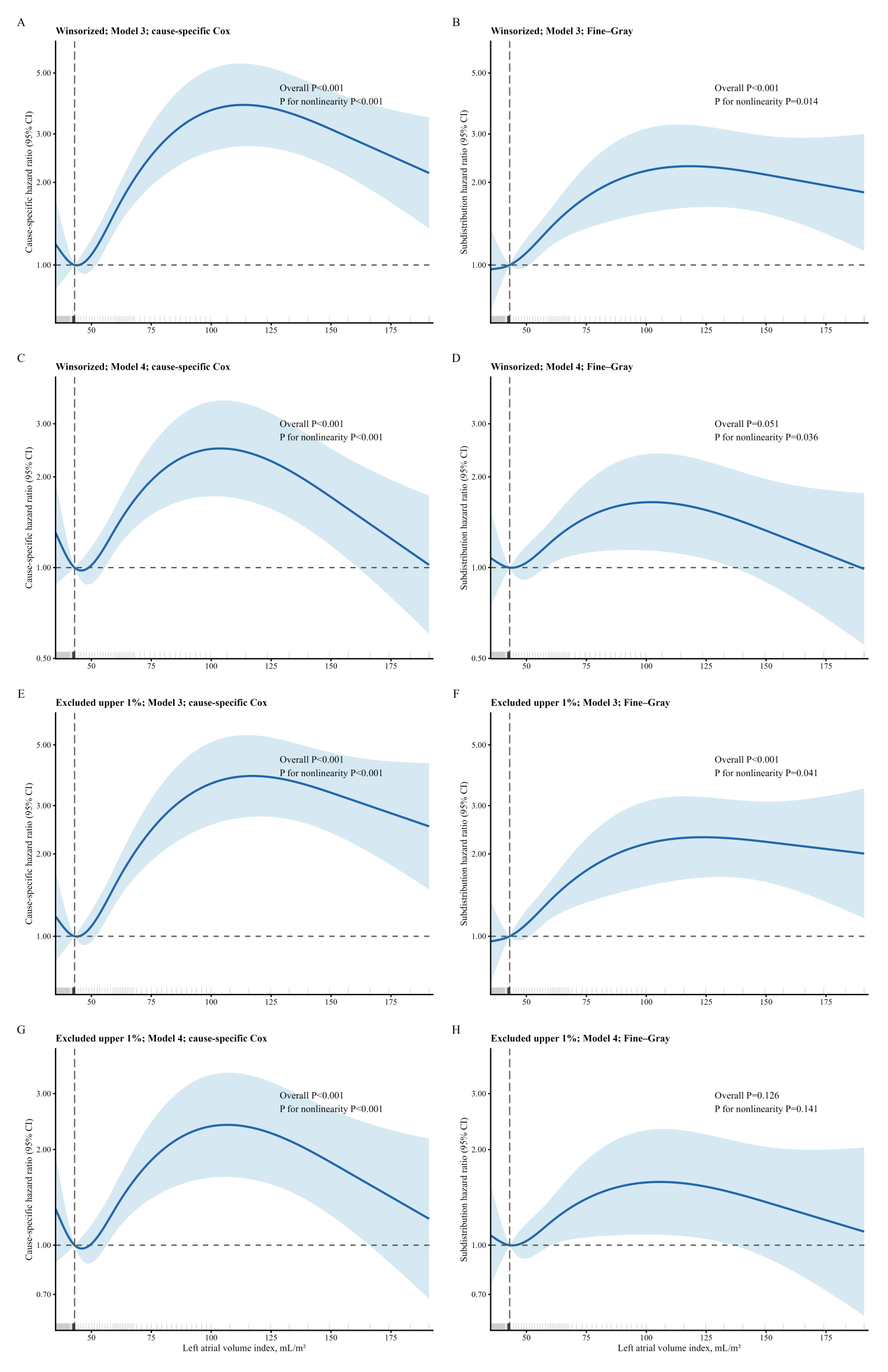

Restricted cubic spline analyses evaluated the association of left atrial volume index with documented incident AF after two approaches to address extreme LAVI values. Panels A–D show analyses in which LAVI was winsorized at the 1st and 99th percentiles; panels E–H show analyses excluding participants with observed LAVI values above the 99th percentile. Panels A, C, E, and G present cause-specific Cox regression results, and panels B, D, F, and H present Fine–Gray competing-risk regression results. Panels A, B, E, and F were adjusted using Model 3, the primary clinical model, which included age, sex, heart rate, left ventricular ejection fraction, left ventricular mass index, diabetes mellitus, renal disease, hypertension, chronic obstructive pulmonary disease, and ischaemic heart disease. Panels C, D, G, and H were adjusted using Model 4, the haemodynamic overlap-adjusted model, which additionally included E/e′ ratio and pulmonary artery systolic pressure. Solid lines represent hazard ratios or subdistribution hazard ratios, and shaded areas indicate 95% confidence intervals. The vertical dashed line indicates the reference LAVI value of 43.0 mL/m², corresponding to the median of the observed pre-imputation distribution. Rug marks along the x-axis show the distribution of LAVI values included in each analysis. Overall and nonlinearity P values are displayed within each panel. Death before documented incident AF was treated as a censoring event in cause-specific Cox regression and as a competing event in Fine–Gray regression. Missing covariate data were handled using multiple imputation by chained equations with 30 imputed data sets, and estimates were pooled using Rubin’s rules. AF, atrial fibrillation; CI, confidence interval; COPD, chronic obstructive pulmonary disease; E/e′, ratio of early transmitral flow velocity to early diastolic mitral annular velocity; LAVI, left atrial volume index; PASP, pulmonary artery systolic pressure.

**Supplementary Figure 3.** Correlations Among Echocardiographic Variables and Assessment of Multicollinearity in the Haemodynamic Overlap-Adjusted Model

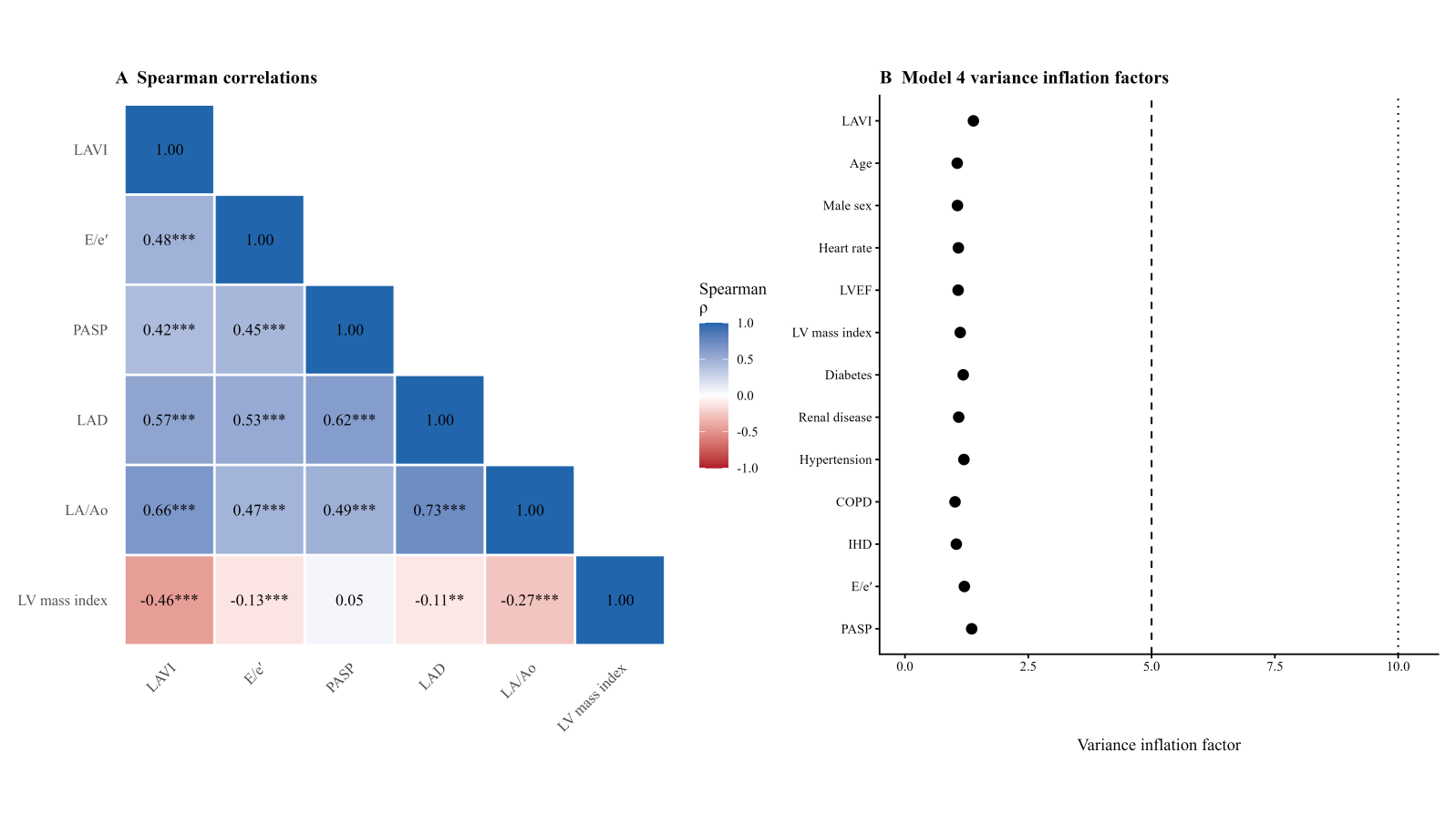

Panel A shows the Spearman correlation matrix for left atrial volume index, E/e′ ratio, pulmonary artery systolic pressure, left atrial diameter, left atrial-to-aortic root ratio, and left ventricular mass index. Values within the cells represent Spearman correlation coefficients (ρ); shading indicates the direction and magnitude of correlation. P values are denoted by asterisks: *P<0.05, **P<0.01, and ***P<0.001. Panel B shows variance inflation factors for the covariates included in Model 4, the haemodynamic overlap-adjusted model: LAVI, age, sex, heart rate, left ventricular ejection fraction, left ventricular mass index, diabetes mellitus, renal disease, hypertension, chronic obstructive pulmonary disease, ischaemic heart disease, E/e′ ratio, and PASP. The dashed and dotted vertical lines indicate variance inflation factor thresholds of 5 and 10, respectively. COPD, chronic obstructive pulmonary disease; IHD, ischaemic heart disease; LA/Ao, left atrial-to-aortic root ratio; LAD, left atrial diameter; LAVI, left atrial volume index; LVEF, left ventricular ejection fraction; PASP, pulmonary artery systolic pressure.

**Supplementary Figure 4.** Scaled Schoenfeld Residual Plots for Assessment of the Proportional Hazards Assumption in Cause-Specific Cox Regression Models

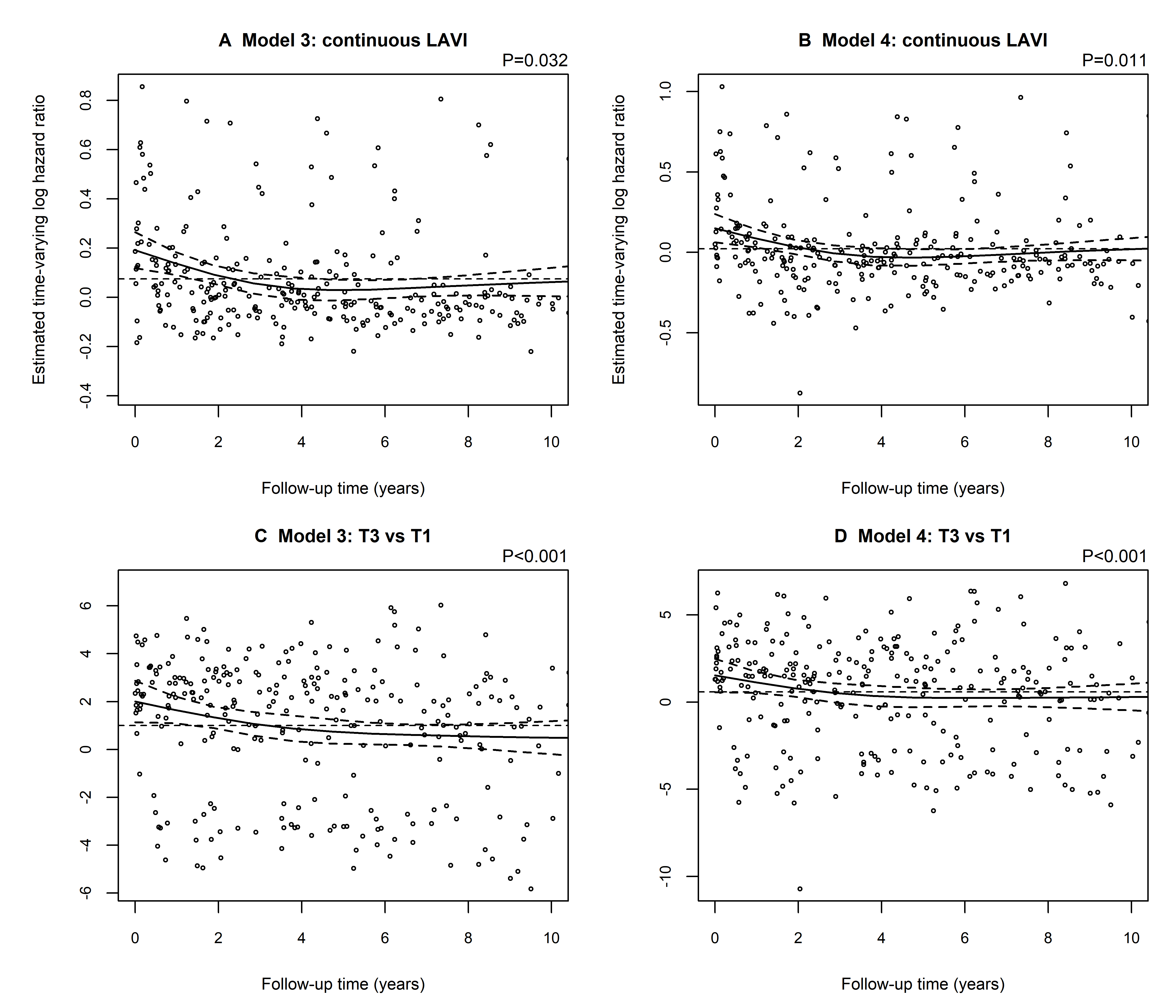

Panels show the estimated time-varying log hazard ratios for continuous LAVI in Model 3 (A) and Model 4 (B), and for the highest versus lowest LAVI tertile in Model 3 (C) and Model 4 (D). Solid curves represent smoothed estimates of the time-varying coefficient, and curved dashed lines represent the corresponding approximate 95% confidence intervals. Horizontal dashed lines indicate the constant log hazard ratio estimated by the corresponding Cox model. Follow-up was displayed through 10 years for graphical presentation. P values correspond to Schoenfeld residual tests for the plotted terms. Diagnostic analyses were performed in the common complete-case sample. Model 3, the primary clinical model, adjusted for age, sex, heart rate, left ventricular ejection fraction, left ventricular mass index, diabetes mellitus, renal disease, hypertension, chronic obstructive pulmonary disease, and ischaemic heart disease. Model 4, the haemodynamic overlap-adjusted model, additionally adjusted for E/e′ ratio and pulmonary artery systolic pressure. AF, atrial fibrillation; E/e′, ratio of early transmitral flow velocity to early diastolic mitral annular velocity; LAVI, left atrial volume index; PASP, pulmonary artery systolic pressure.
